# A systematic review and meta-analysis comparing the diagnostic accuracy tests of COVID-19

**DOI:** 10.1101/2022.11.29.22282895

**Authors:** Juan Jeferson Vilca-Alosilla, Mayron Antonio Candia-Puma, Katiusca Coronel-Monje, Luis Daniel Goyzueta-Mamani, Alexsandro Sobreira Galdino, Ricardo Andrez Machado-de-Ávila, Rodolfo Cordeiro Giunchetti, Eduardo Antonio Ferraz Coelho, Miguel Angel Chávez-Fumagalli

**Affiliations:** Computational Biology and Chemistry Research Group, Vicerrectorado de Investigación, Universidad Católica de Santa María, Arequipa 04000, Peru; Facultad de Ciencias Farmacéuticas, Bioquímicas y Biotecnológicas, Universidad Católica de Santa María, Arequipa 04000, Peru; Sustainable Innovative Biomaterials Department, Le Qara Research Center, Arequipa 04000, Peru; Laboratório de Biotecnologia de Microrganismos, Universidade Federal São João Del-Rei, Divinópolis 35501-296, MG, Brazil; Programa de Pós-Graduação em Ciências da Saúde, Universidade do Extremo Sul Catarinense, Criciúma 88806-000, SC, Brazil; Laboratório de Biologia das Interações Celulares, Instituto de Ciências Biológicas, Universidade Federal de Minas Gerais, Belo Horizonte 31270-901, MG, Brazil; Instituto Nacional de Ciência e Tecnologia em Doenças Tropicais, INCT-DT, Salvador 40015-970, BA, Brazil; Programa de Pós-Graduação em Ciências da Saúde: Infectologia e Medicina Tropical, Faculdade de Medicina, Universidade Federal de Minas Gerais, Belo Horizonte 31270-901, MG, Brazil; Departamento de Patologia Clínica, COLTEC, Universidade Federal de Minas Gerais, Belo Horizonte 31270-901, MG, Brazil

**Keywords:** SARS-CoV-2, Diagnostic Tests, Meta-analysis, Systematic Review, Sensitivity, Specificity

## Abstract

In this work, we report a systematic review and meta-analysis that seeks to analyze the accuracy of diagnostic tests for coronavirus disease 2019 (COVID-19). The objective of this article is to detail the scientific findings based on diagnostic tests of the last years when the pandemic caused by severe acute respiratory syndrome coronavirus 2 (SARS-CoV-2) occurred. Searches for published studies were carried out in the PubMed database between the years 2020 and 2021 for the diagnosis of COVID-19. Ninety-nine scientific articles that met the criteria were examined and accepted in the meta-analysis, and the diagnostic accuracy was evaluated through specificity and sensitivity. Molecular tests [Reverse transcription polymerase chain reaction (RT-PCR), reverse transcription loop-mediated isothermal amplification (RT-LAMP), and clustered regularly interspaced short palindromic repeats (CRISPR)] showed better performance in terms of sensitivity and specificity when compared to serological tests [Enzyme-linked immunosorbent assay (ELISA), chemiluminescence immunoassay (CLIA), lateral flow immunoassay (LFIA), chemiluminescent microparticle immunoassays (CMIA), and Fluorescence immunoassay (FIA)], which showed higher specificity, mainly for the detection of IgG antibodies; however, they showed sensitivity <90%. In addition, the antiviral neutralization bioassay (ANB) diagnostic test demonstrated high potential for the diagnosis of COVID-19, since it obtained the highest area under the curve restricted to the false-positive rates (AUC_FPR_) of 0.984. It is settled that the different diagnostic tests have been efficiently adapted for the detection of SARS-CoV-2; however, their performance still needs to be optimized to control future outbreaks of COVID-19, which will also serve to help the control of future infectious agents.

## Introduction

Coronavirus disease 2019 (COVID-19), caused by severe acute respiratory syndrome coronavirus 2 (SARS-CoV-2), was first reported at the end of 2019 in Wuhan, Hubei province, China [1]. On 11 March 2020, the World Health Organization (WHO) declared COVID-19 a pandemic, due to the high levels of spread worldwide [2]. The virus is transmitted by direct contact with an infected person, by the expulsion of droplets and small particles when breathing, speaking, or coughing, or by drops of saliva with the virus being deposited on surfaces and/or objects, which are transmitted through touch [3,4]. According to the WHO, by September 2022, around 600 million confirmed cases of COVID-19 have been reported worldwide, including more than 6 million deaths [5]. Likewise, the transmission of the virus by asymptomatic people is a major concern for community spread [6], where a study indicates that 35.1% of patients with COVID-19 did not present symptoms [7]; in this context, so an early diagnosis of infection would allow the rapid spread of the virus to be controlled [8]. It was estimated that a single symptomatic COVID-19 infection would have an average direct medical cost during infection condition of US$3 045, which would increase to $14 366 per hospitalization [9], making COVID-19 one of the greatest and most significant global health crises crisis in human history.

The pathogenicity of SARS-CoV-2 is fundamentally related to the interaction of the virus S protein and host membrane receptor angiotensin-converting enzyme 2 (ACE2) [10]. COVID-19 affects not only the respiratory system but also other organs, such as the liver and kidneys [11]. Generally, the clinical manifestations of COVID-19 are diverse, while the most prevalent symptoms are fever, cough, dyspnea, malaise, fatigue, neurological symptoms, dermatological manifestations, anorexia, myalgia, sneezing, sore throat, rhinitis, goosebumps, headache, chest pain, and diarrhea [12,13]. The symptoms that SARS-CoV-2 induces in several patients are mild, but may also be related to an accelerated onset of generalized infection in the lungs that is complicated by acute respiratory distress syndrome (ARDS) [14,15], due to an unregulated inflammatory/immune system and cytokine storm [16]. In addition, a series of possible late complications due to COVID-19 infection has been pointed out, including venous thromboembolism, arterial thrombosis, pulmonary fibrosis, cardiac thrombosis and inflammation, cerebrovascular accident, mental fog, dermatological complications, and overall mood dysfunctions [17].

Control strategies against the spread of COVID-19 in many countries initially focused on complete or partial lockdowns. Still, these measures were not as effective due to the rapid increase in the number of hospitalized infected people [16]. Several vaccines have been licensed to reduce the incidence of hospitalization and death; despite this vaccination coverage remains insufficient [18,19], and many of them may not be effective against new virus variants [20]. Symptomatic treatment instead of curative treatment has been the fundamental tool for the treatment of critically ill patients through ventilatory support in the intensive care unit [21]. Other potential therapeutic strategies include antiviral, antibiotic, and immunomodulatory therapies [22]. For this reason, diagnostic tools continue to play a crucial role in the battle against COVID-19, allowing rapid implementation of control measures to suppress SARS-CoV-2 transmission through case identification, contact tracing, and isolation of positive cases [23,24].

At the beginning of the COVID-19 pandemic, no efficient diagnostic test was available for patients who became severely ill, and the diagnosis was based mainly on the patient’s clinical manifestations and exposure history. With the SARS-CoV-2 genome sequenced, WHO produced the protocol for the molecular diagnosis of COVID-19 based on real-time RT-PCR, allowing the development of commercial diagnostic kits [25]. Real-time RT-PCR is the current reference laboratory test, considered the “*gold standard*” for its high sensitivity and specificity [26]. However, despite the high diagnostic accuracy, the high cost, the need for ribonucleic acid (RNA) extraction, the availability of specialized raw materials, and the relatively long execution time, makes it is difficult to be applied on a large scale [27,28]. On the other side, serological tests are reliable, simple, and inexpensive techniques that allow direct and indirect detection of infections; however, they detect the presence of antibodies as a marker of past infection [29]. Serologic assays such as Enzyme-Linked Immunosorbent Assay (ELISA), Chemiluminescent Immunoassay (CLIA), and Lateral Flow Immunochromatographic Assay (LFIA) are used as diagnostic tools, often for the detection of IgA, IgG, and IgM antibodies from the patient’s serum or plasma, which are directed against the spike (S) and the nucleocapsid (N) proteins of SARS-CoV-2 [30]. The timing of immunoglobulin production (from 4 days after the onset of symptoms to 10-14 days) limits its applicability in acute phase diagnosis, however, by detecting IgM and IgA that is rapidly formed in response to infection, may represent a tool that can help diagnose COVID-19, as well as significantly increase diagnostic sensitivity by combining serological tests with molecular tests [31,32]. COVID-19 antigen tests allow the diagnosis of active infection by detecting the proteins of the SARS-CoV-2 virus in different types of samples. They are available as single-use, lateral flow, rapid antigen detection diagnostic tests, which can be read visually or processed and read with a small handheld device. Both can be performed outside a laboratory and provide results in 15 to 20 minutes. These tests can be produced faster and applied on a larger scale. While these tests can be very specific, they are generally not as sensitive as molecular tests [33–35].

The present study aims to systematically review and summarize the available literature on the diagnostic accuracy of COVID-19 diagnostic tests. In this sense, a systematic review of the literature was carried out and the results were analyzed through a meta-analysis based on the techniques developed and used for the diagnosis of COVID-19. The diagnostic techniques analyzed were reverse transcription–polymerase chain reaction (RT-PCR), reverse transcriptase loop-mediated isothermal amplification (RT-LAMP), clustered regularly interspaced short palindromic repeats (CRISPR), microarrays (MA), next-generation sequencing (NGS), enzyme-linked immunosorbent assay (ELISA), antiviral neutralization bioassay (ANB), biosensors (BS), chemiluminescence immunoassay (CLIA), lateral flow immunoassay (LFIA), chemiluminescent microparticle immunoassay (CMIA), electrochemiluminescence immunoassay (ECLIA) and fluorescence immunoassay (FIA). Therefore, we hope that the data generated will help identify the most effective diagnostic techniques against new pathogens such as SARS-CoV-2 and its variants.

## Methods

### Search strategy

This systematic review was based on the PRISMA (Preferred Reporting Items for Systematic Reviews and Meta-Analyses) technique [36], and this systematic review protocol has been registered on INPLASY (https://inplasy.com/inplasy-2022-9-0132), while the registration number is INPLASY202290132. The search was carried out and conducted until 10 June 2022 in the PubMed database (https://pubmed.ncbi.nlm.nih.gov/). PubMed is one of the most widely used search engines for biomedical literature, developed and supported by the US NLM/National Center for Biotechnology Information (NCBI) [37]. The search for the terms associated in the literature with the diagnosis of COVID-19 was carried out using the MeSH term “COVID-19”, the results were shown in a co-occurrence network map of MeSH terms in the VOSviewer software (version 1.6.18) [38]. The clusters in the network map were analyzed to choose relevant terms associated with COVID-19 diagnostic techniques. Additionally, a second search was developed with each MeSH term obtained in the cluster analysis, relating the MeSH term “sensitivity and specificity”, which are normally typically considered a parameter for the evaluation of the diagnostic accuracy of a test [39], and the MeSH term “COVID-19”.

### Search strategy, eligibility criteria, and data extraction

The studies included in the meta-analysis were chosen in three stages. In the first stage duplicate articles, articles in languages other than English, reviews, and meta-analyses were excluded. In the second stage of the screening stage, the titles and abstracts of the articles found through the search strategy were analyzed. Finally, in the eligibility stage, the complete studies with high relevance were obtained, and they were isolated from the articles with a title or abstract that did not provide sufficient data to be considered within the meta-analysis. The information extracted from each selected study included the diagnostic technique, the number, type, and clinical characteristics of the patients with COVID-19, and healthy controls. All studies evaluating diagnostic accuracy using sensitivity and specificity parameters have been included. Likewise, the data related to the geographical distribution, the number of studies related by country, and the frequency of the diagnostic techniques used were extracted. Studies lack data of Studies with unclear or lacking data regarding the sensitivity and specificity of COVID-19 diagnostic tests were excluded from further analysis.

### Statistical analysis

Results were entered into Microsoft Excel (version 10.0, Microsoft Corporation, Redmond, WA, USA) spreadsheets and analyzed in the R programming environment (version 4.2.1) using the package *“mada”* (version 0.5.11) https://cran.r-project.org/web/packages/mada/index.html; which employs a hierarchical model that accounts for within and between-study (heterogeneity) and the correlation between sensitivity and specificity [40]. Initially, the number of true negatives (TP), false negatives (FN), true positives (TP), and false positives (FP) were analyzed separately for each diagnostic technique; while the evaluation of sensitivity (Se) and specificity (Sp) made it possible to determine the diagnostic performance. Additionally, the positive likelihood ratio (LR+) expresses the ratio between the probability of expecting a positive test in a patient and the probability of expecting a positive test in a patient without the disease [41]; the negative likelihood ratio (LR−), which expresses the probability that a patient will test negative among people with the disease and the probability that a patient will test negative among people without disease; and the diagnostic likelihood ratio (DOR), which is the odds ratio of the positivity of a diagnostic test result in the diseased population relative to the non-diseased population [42]; and the 95% confidence interval (CI) were determined. Summary receiver operating characteristic (sROC) curves were fitted, according to the parameters of the *“Reitsma”* model of the *“mada”* package, and were used to compare the diagnostic accuracy of COVID-19 diagnostic techniques [43]. The confidence level for all calculations was set to 95%, using a continuity correction of 0.5 if pertinent.

## Results

### Data sources and study selection

In this study, a systematic review followed by meta-analysis was performed to measure the accuracy of diagnostic tests for COVID-19. A flowchart of the study strategy was prepared and presented (Figure 1). To this end, a search was accomplished in the PubMed database with the MeSH terms “COVID-19”, and a co-occurrence network map of MeSH terms was developed; Through the search, 981 scientific articles were obtained between the years 2020 and 2021. The minimum number of occurrences of keywords was set at a value of five, and a network map with 2.518 keywords was generated (Figure 2A). The formation of five main clusters was found in the analysis of the network map, in the cluster related to diagnostic techniques (purple color) terms such as *“Reverse Transcription–Polymerase Chain Reaction”, “Reverse Transcriptase Loop-Mediated Isothermal Amplification”, “Clustered Regularly Interspaced Short Palindromic Repeats”, “Microarrays”, “Next-Generation Sequencing”, “Enzyme-Linked Immunosorbent Assay”, “Antiviral Neutralization Bioassay”, “Biosensor”* and *“Immunoassay”.* In addition, terms such as *”COVID-19”, “SARS-COV-2”, “adult”, China”, “disease outbreaks middle-aged”,* and *”female”* were common denominators (Figure 2A).

**Figure 1.**
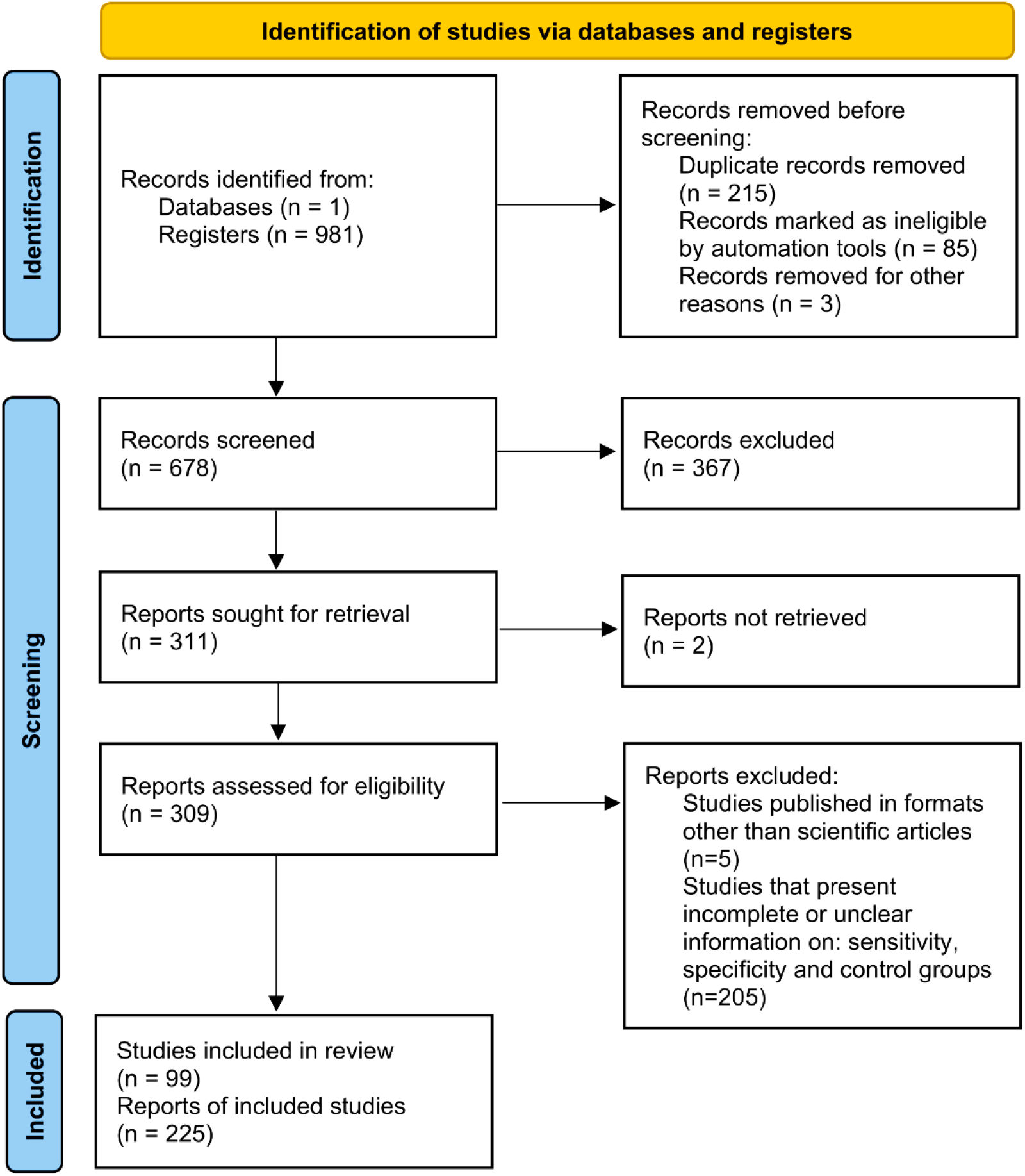
A systematic review and meta-analysis flow diagram of the study selection process.

**Figure 2.**
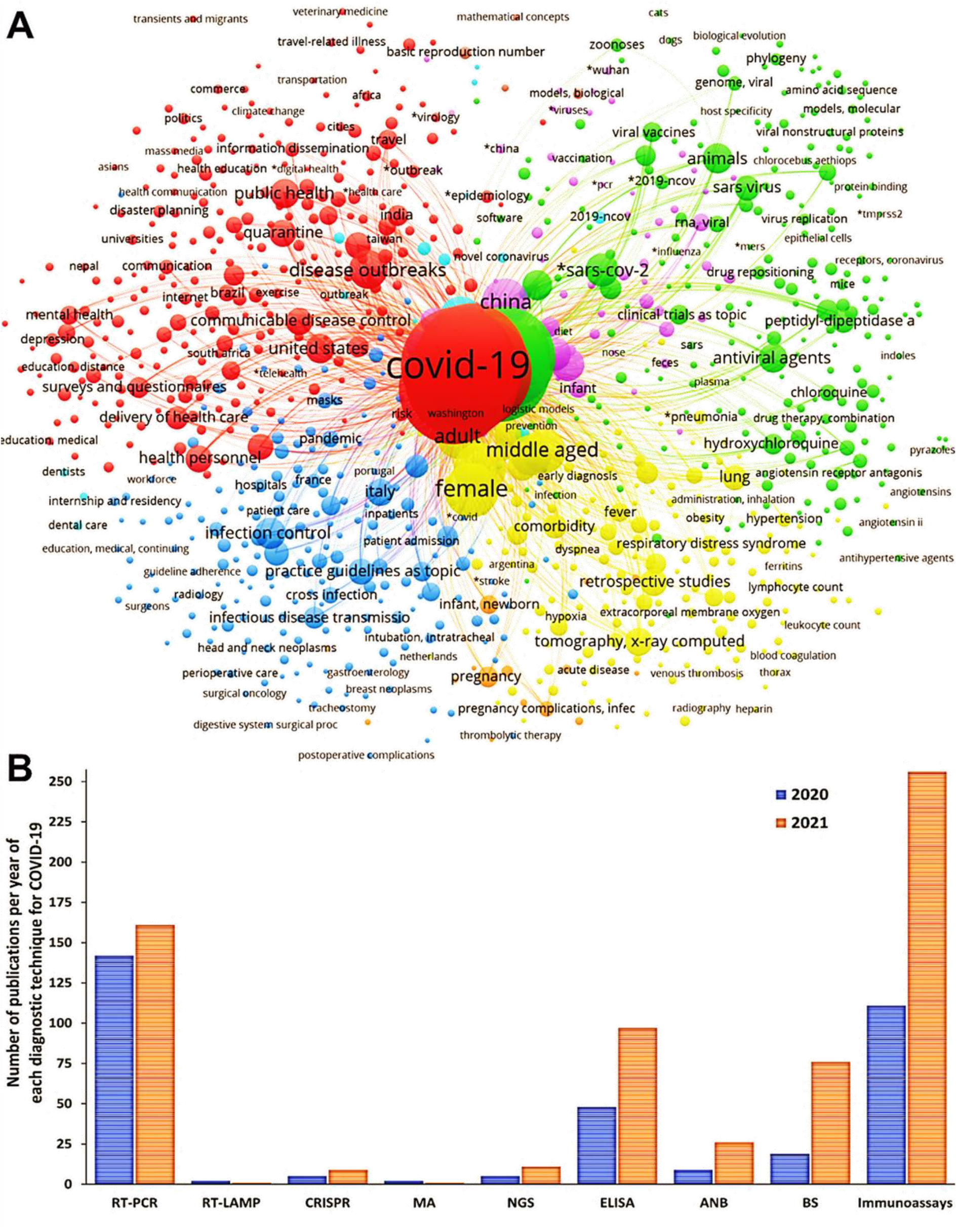
Selected articles using the PubMed database for the different diagnostic techniques using MeSH terms. A) Network map built by VOSviewer based on the co-occurrence of MeSH terms. B) Number of articles found with the search for each diagnostic test, considered from cluster analysis.

A second search was performed in the PubMed database with the terms found in the first analysis. The new terms were associated with the terms “*COVID-19*” and “*Sensitivity and Specificity*”; generating the new search strings: (Covid 19[MeSH Terms]) AND (sensitivity and specificity[MeSH Terms]) AND (RT-PCR[MeSH Terms]) for RT-PCR; (Covid 19[MeSH Terms]) AND (sensitivity and specificity[MeSH Terms]) AND (RT-LAMP assay[MeSH Terms]) for RT-LAMP; (Covid 19[MeSH Terms]) AND (sensitivity and specificity[MeSH Terms]) AND (CRISPR[MeSH Terms]) for CRISPR; (Covid 19[MeSH Terms]) AND (sensitivity and specificity[MeSH Terms]) AND (Microarray Analysis[MeSH Terms]) for MA; (Covid 19[MeSH Terms]) AND (sensitivity and specificity[MeSH Terms]) AND (Next generation sequencing[MeSH Terms]) for NGS; (Covid 19[MeSH Terms]) AND (sensitivity and specificity[MeSH Terms]) AND (ELISA[MeSH Terms]) for ELISA; (Covid 19[MeSH Terms]) AND (sensitivity and specificity[MeSH Terms]) AND (Neutralization Tests[MeSH Terms]) for ANB; (Covid 19[MeSH Terms]) AND (sensitivity and specificity[MeSH Terms]) AND (Biosensing Technique[MeSH Terms]) for BS; and (Covid 19[MeSH Terms]) AND (sensitivity and specificity[MeSH Terms]) AND (Immunoassay[MeSH Terms]) for immunoassays.

The number of studies chosen for RT-PCR, RT-LAMP, CRISPR, MA, NGS, ELISA, ANB, BS, and immunoassays were: 303, 3, 14, 3, 16, 145, 35, 95, and 367, respectively (Figure 2B). The three-step eligibility criterion allowed 303, 369, and 215 articles to be excluded, from the criteria for identification, screening, and eligibility, respectively. Therefore, 99 articles were selected for meta-analysis (Figure 3). It is observed that in most studies, the diagnostic techniques used were immunoassays (CLIA, LFIA, CMIA, ECLIA, and FIA) (Figure 3A). Additionally, China, the United States of America, and India are the countries that have carried out a higher number of studies related to diagnostic tests for COVID-19 (Figures 3B and 3C).

**Figure 3.**
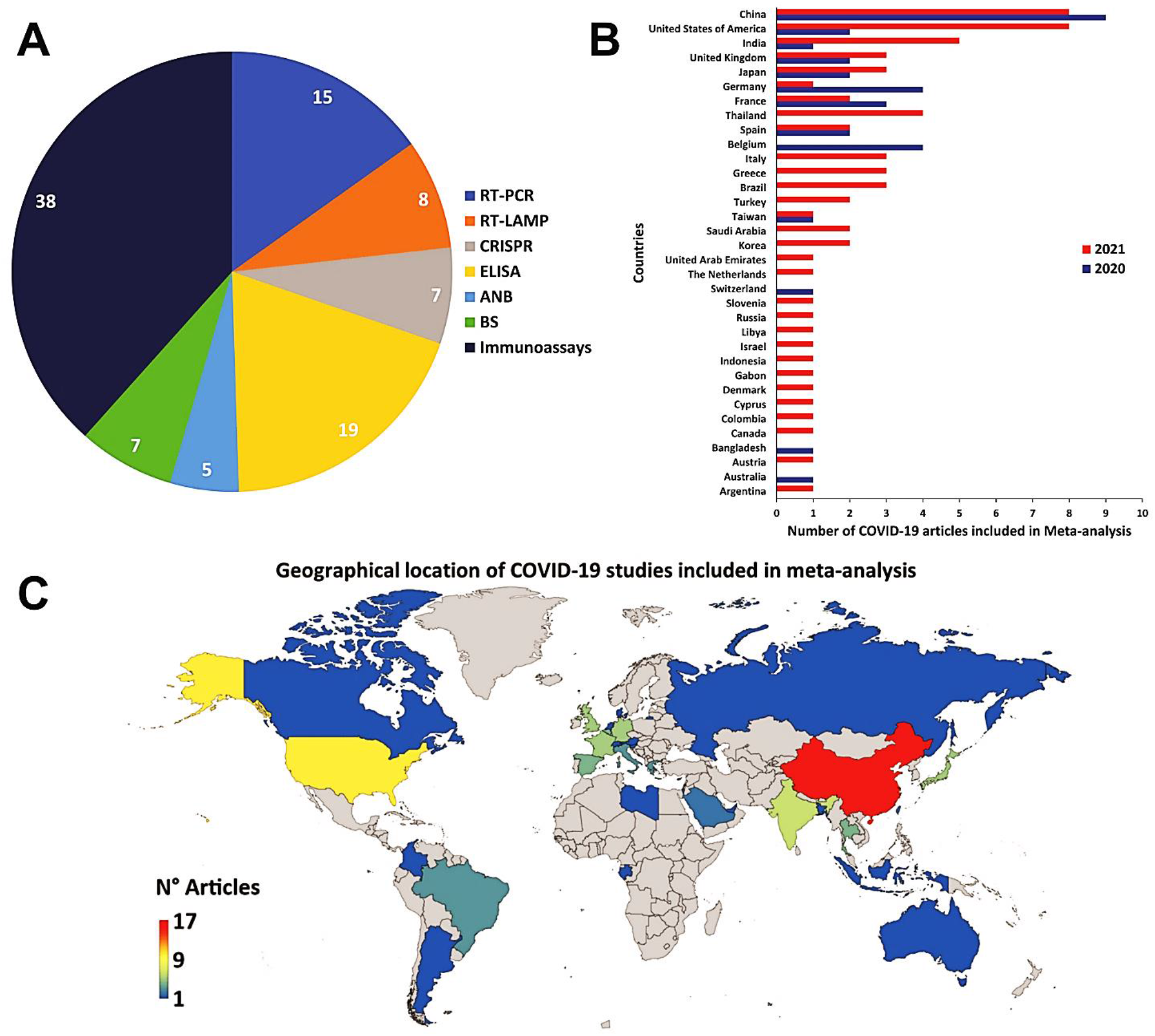
The geographical location of COVID-19 studies. A) The pie chart shows the type of diagnostic tests used in the COVID-19 studies for the meta-analysis. B) The bar graph shows the number of COVID-19 studies, included in the meta-analysis, carried out by different countries. C) Demographic representation of COVID-19 studies, included in the meta-analysis, worldwide (lower-blue to upper-red numbers)

### Meta-analysis of the diagnostic techniques for COVID-19

#### Reverse transcription–polymerase chain reaction

Fifteen studies based on the RT-PCR technique were selected [44–58], in which a total of 6,902 subjects were studied. Sensitivity ranged from 36.8% to 99.2%, with a median of 94.5%, 95%CI (85.1, 98.0), while the test for equality of sensitivities presented a χ^2^ = 577.02, df = 38, p-value = 2e^-16^. Specificity ranged from 79.3 to 99.8%, with a median of 98.4%, 95%CI (86.7, 99.8); the test for equality of specificities showed χ^2^ = 142.18, df = 38, p-value = 5.96e^-14^. A negative correlation between sensitivities and false positive rates is shown r = –0.188, 95%CI (–0.476, 0.135). Additionally, results regarding LR+ {median 45.00, 95%CI (4.06, 563.27)}, LR− {median 0.06, 95%CI (0.02, 0.18)} and DOR {median 609.00, 95%CI (54.53, 8485.95)}. The analyzed diagnostic performance is summarized in Figure 4 and Supplementary Figure 1.

**Figure 4.**
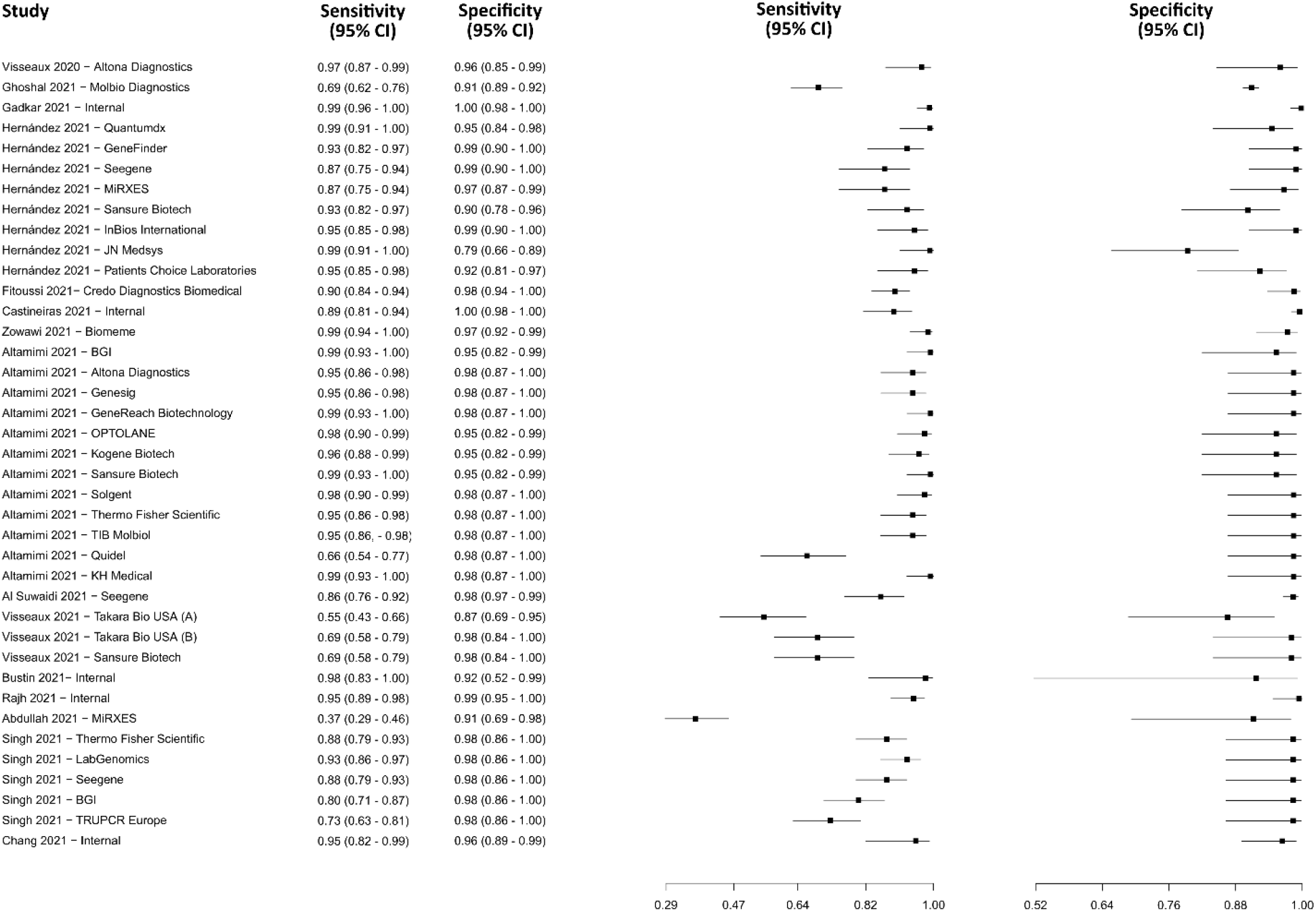
Data analysis and paired forest plot of the sensitivity and specificity of the reverse transcription–polymerase chain reaction (RT-PCR) in the diagnosis of COVID-19. Sensitivity and specificity are reported as a mean (95% confidence limits). The forest plot represents the estimated sensitivity and specificity (black squares) and their 95% confidence limits (horizontal black line) [44–58].

#### Reverse transcriptase loop-mediated isothermal amplification

Seven studies were selected using the RT-LAMP technique [59–65]. A total of 1,806 subjects were studied. Sensitivity ranged from 74.7 to 98.8%, with a median of 91.9%, 95%CI (80.0, 97.0); while the test for equality of sensitivities showed: χ^2^ = 30.09, df =7, p-value = 9.12e^-05^. Specificity ranged from 88.1 to 99.6%, with a median of 98.8%, 95%CI (90.0, 100.0); while the test for equality of specificities presented χ^2^ = 34.71, df = 7, p-value = 1.27e^-05^. The correlation between sensitivities and false positive rates was analyzed a negative result is shown r = 0.313, 95%CI (-0.502, 0.834). In addition, results regarding LR+ {median 69.51, 95%CI (4.88, 755.24)}, LR− {median 0.09, 95%CI (0.03, 0.23)} and DOR {median 801.74, 95%CI (48.36, 10044.33)} are displayed. The analyzed diagnostic performance is summarized in Figure 5 and Supplementary Figure 2.

**Figure 5.**
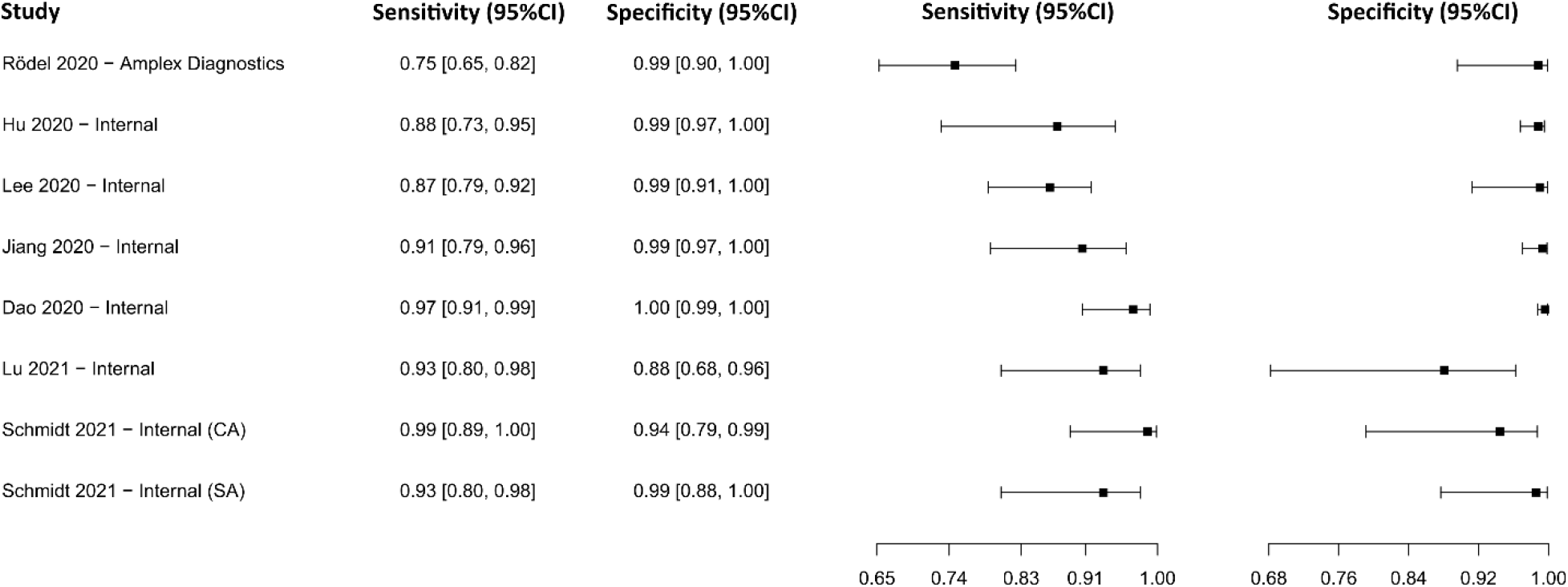
Data analysis and paired forest plot of the sensitivity and specificity of the reverse transcriptase loop-mediated isothermal amplification (RT-LAMP) in the diagnosis of COVID-19. Sensitivity and specificity are reported as a mean (95% confidence limits). The forest plot represents the estimated sensitivity and specificity (black squares) and their 95% confidence limits (horizontal black line) [59–65].

#### Clustered regularly interspaced short palindromic repeats

The analysis identified 14 published studies that used CRISPR as a diagnostic tool. After analysis, only 7 seven studies [66–72] were selected. A total of 1,201 subjects were studied. Sensitivity ranged from 67.0 to 99.5%, with a median of 94.4%, and 95%CI (84.0, 99.0). Test for equality of sensitivities analysis showed: χ^2^ = 80.26, df = 10, p-value = 4.47e^-13^. Specificity ranged from 83.6 to 99.6%, with a median of 98.6%, 95%CI (93.0, 100.0); while the test for equality of specificities: χ^2^ = 55.37, df = 10, p-value = 2.69e^-08^. Also, a negative correlation between sensitivities and false positive rates is shown r = 0.328, 95%CI (–0.339, 0.775). In addition, results regarding LR+ {median 70.33, 95%CI (6.80, 898.96)}, LR− {median 0.06, 95%CI (0.01, 0.20)} and DOR {median 1357, 95%CI (37.27, 29912.15)} are displayed. The diagnostic performance of the selected studies is summarized in Figure 6 and Supplementary Figure 3.

**Figure 6.**
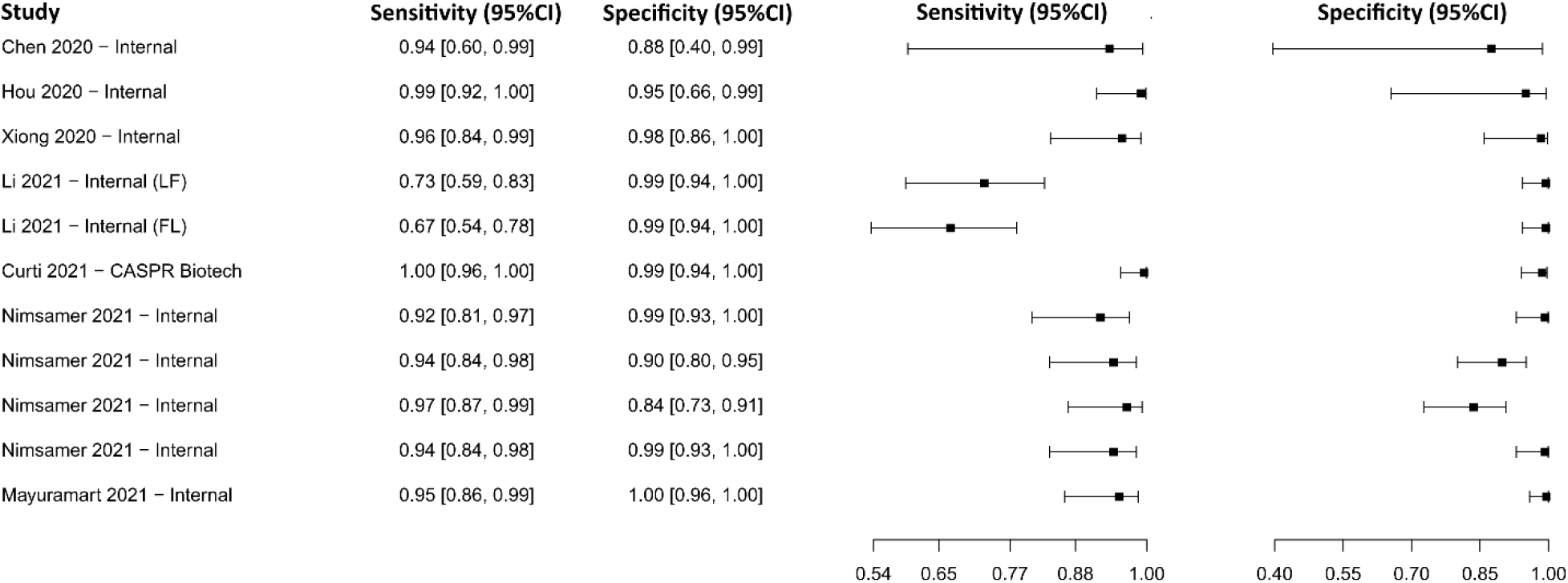
Data analysis and paired forest plot of the sensitivity and specificity of the clustered regularly interspaced short palindromic repeats (CRISPR) in the diagnosis of COVID-19. Sensitivity and specificity are reported as a mean (95% confidence limits). The forest plot represents the estimated sensitivity and specificity (black squares) and their 95% confidence limits (horizontal black line) [66–72].

#### Enzyme-linked immunosorbent assay for IgG

Sixteen studies were selected using IgG-detecting ELISA as a diagnostic technique [73–88]. A total of 9,221 subjects were studied. Sensitivity ranged from 61.6 to 99.0%, with a median of 88.3%, 95%CI (80.3, 93.5), while the test for equality of sensitivities presented a χ^2^ = 151.80, df = 22, p-value = 2e^-16^. Specificity ranged from 81.3 to 99.7%, with a median of 97.4%, 95%CI (89.6, 98.9); the test for equality of specificities showed χ^2^ = 135.69, df = 22, p-value = 2e^-16^. A negative correlation between sensitivities and false positive rates is shown r = 0.166, 95%CI (–0.264, 0.541). In addition, the results regarding LR+ {median 29.99, 95%CI (5.78, 80.78)}, LR− {median 0.12, 95%CI (0.06, 0.23)} and DOR {median 333.00, 95%CI (37.03, 1288.51)}. The analyzed diagnostic performance is summarized in Figure 7 and Supplementary Figure 4.

**Figure 7.**
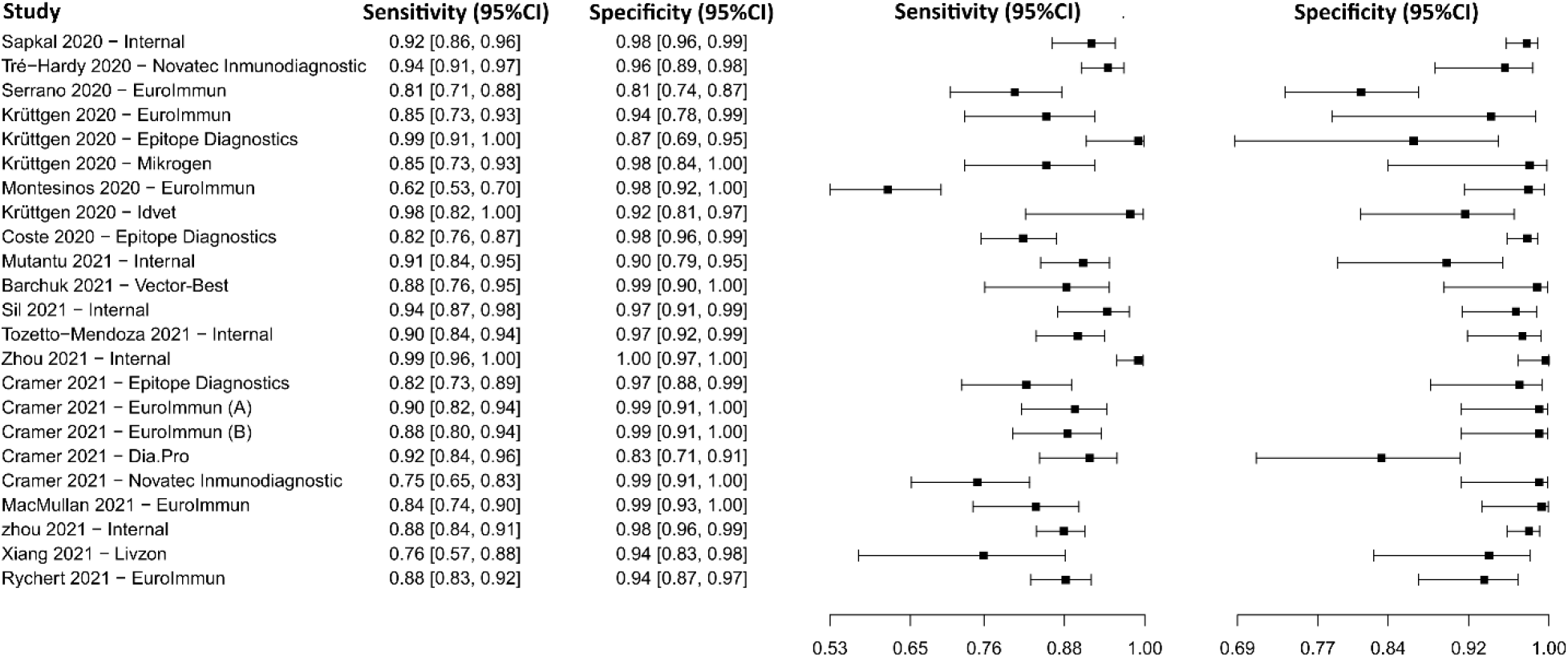
Data analysis and paired forest plot of the sensitivity and specificity of the enzyme-linked immunosorbent assay (ELISA) for IgG in the diagnosis of COVID-19. Sensitivity and specificity are reported as a mean (95% confidence limits). The forest plot represents the estimated sensitivity and specificity (black squares) and their 95% confidence limits (horizontal black line) [73–88].

#### Enzyme-linked immunosorbent assay for IgM

Five studies were selected using IgM-detecting ELISA as a diagnostic tool [74,84,87,89,90], in which a total of 1,585 subjects were studied. Sensitivity ranged from 46.9% to 99.7%, with a median of 73.1%, 95%CI (57.3, 78.8), while the test for equality of sensitivities presented a χ^2^ = 150.59, df = 4, p-value = 2e^-16^. Specificity ranged from 89.3 to 99.8%, with a median of 98.1%, 95%CI (92.3, 99.6); the test for equality of specificities showed χ^2^ = 28,12, df = 4, p-value = 1.18e^-05^. A negative correlation between sensitivities and false positive rates is shown r = 0.446, 95%CI (–0.719, 0.953). Additionally, results regarding LR+ {median 25.90, 95%CI (9.29, 127.19)}, LR− {median 0.27, 95%CI (0.21, 0.53)} and DOR {median 546.65, 95%CI (33.36, 8956.27)}. The analyzed diagnostic performance is summarized in Figure 8 and Supplementary Figure 5.

**Figure 8.**
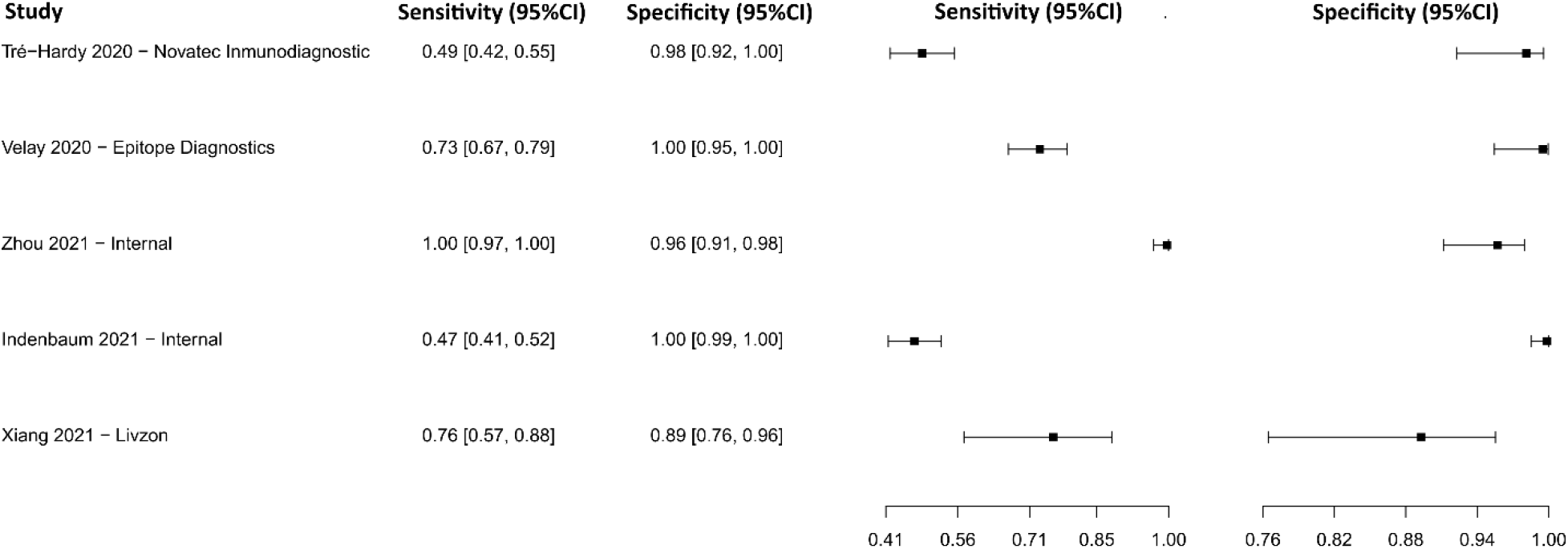
Data analysis and paired forest plot of the sensitivity and specificity of the enzyme-linked immunosorbent assay (ELISA) for IgM in the diagnosis of COVID-19. Sensitivity and specificity are reported as a mean (95% confidence limits). The forest plot represents the estimated sensitivity and specificity (black squares) and their 95% confidence limits (horizontal black line) [74,84,87,89,90].

#### Enzyme-linked immunosorbent assay for IgA

Five studies were selected using IgA-detecting ELISA as a diagnostic technique [74,75,77,89,90]. A total of 1,632 subjects were studied. Sensitivity ranged from 79.8 to 92.4%, with a median of 83.7%, 95%CI (77.9, 88.8);), while the test for equality of sensitivities showed: χ^2^ = 13.71, df =4, p-value = 8.25e^-03^. Specificity ranged from 85.6 to 99.6%, with a median of 98.0%, 95%CI (92.3, 99.1); while the test for equality of specificities presented χ^2^ = 34.46, df = 4, p-value = 6.00e^-07^. The correlation between sensitivities and false positive rates was analyzed a negative result is shown r = -0.448, 95%CI (-0.953, 0.718). In addition, the results regarding LR+ {median 39.92, 95%CI (9.80, 85.62)}, LR− {median 0.18, 95%CI (0.13, 0.24)} and DOR {median 194.04, 95%CI (85.07, 442.62)} are displayed. The analyzed diagnostic performances are summarized in Figure 9 and Supplementary Figure 6.

**Figure 9.**
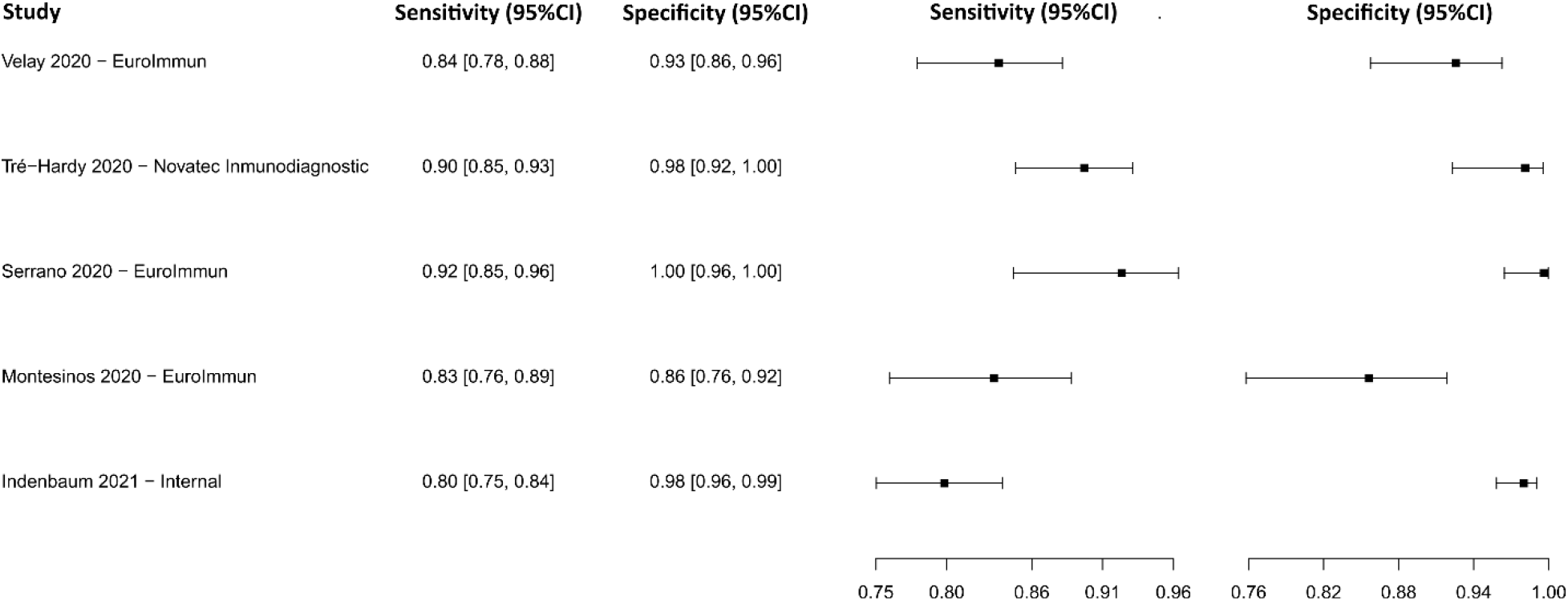
Data analysis and paired forest plot of the sensitivity and specificity of the enzyme-linked immunosorbent assay (ELISA) for IgA in the diagnosis of COVID-19. Sensitivity and specificity are reported as a mean (95% confidence limits). The forest plot represents the estimated sensitivity and specificity (black squares) and their 95% confidence limits (horizontal black line) [74,75,77,89,90].

#### Antiviral neutralization bioassay

The analysis identified 40 studies that used ANB as a diagnostic technique for COVID-19. After analysis, only 5 five studies [91–95] were selected. A total of 1,567 subjects were studied. Sensitivity ranged from 90.2 to 98.8%, with a median of 95.6%, and 95%CI (90.0, 98.0). Test for equality of sensitivities analysis showed: χ^2^ = 19.18, df = 4, p-value = 7.23e^-04^. The specificity of the studies ranged from 98.7 to 99.8%, with a median of 99.5%, 95%CI (96.0, 100.0); while the test for equality of specificities: χ^2^ = 3.21, df = 4, p-value = 0.52. Also, a negative correlation between sensitivities and false positive rates is shown r = -0.055, 95%CI (–0.894, 0.869). In addition, results regarding LR+ {median 185.17, 95%CI (22.71, 2950.41)}, LR− {median 0.04, 95%CI (0.02, 0.10)} and DOR {median 6332.92, 95%CI (458.41, 56606.18)} are displayed. The diagnostic performance of the selected studies is summarized in Figure 10 and Supplementary Figure 7.

**Figure 10.**
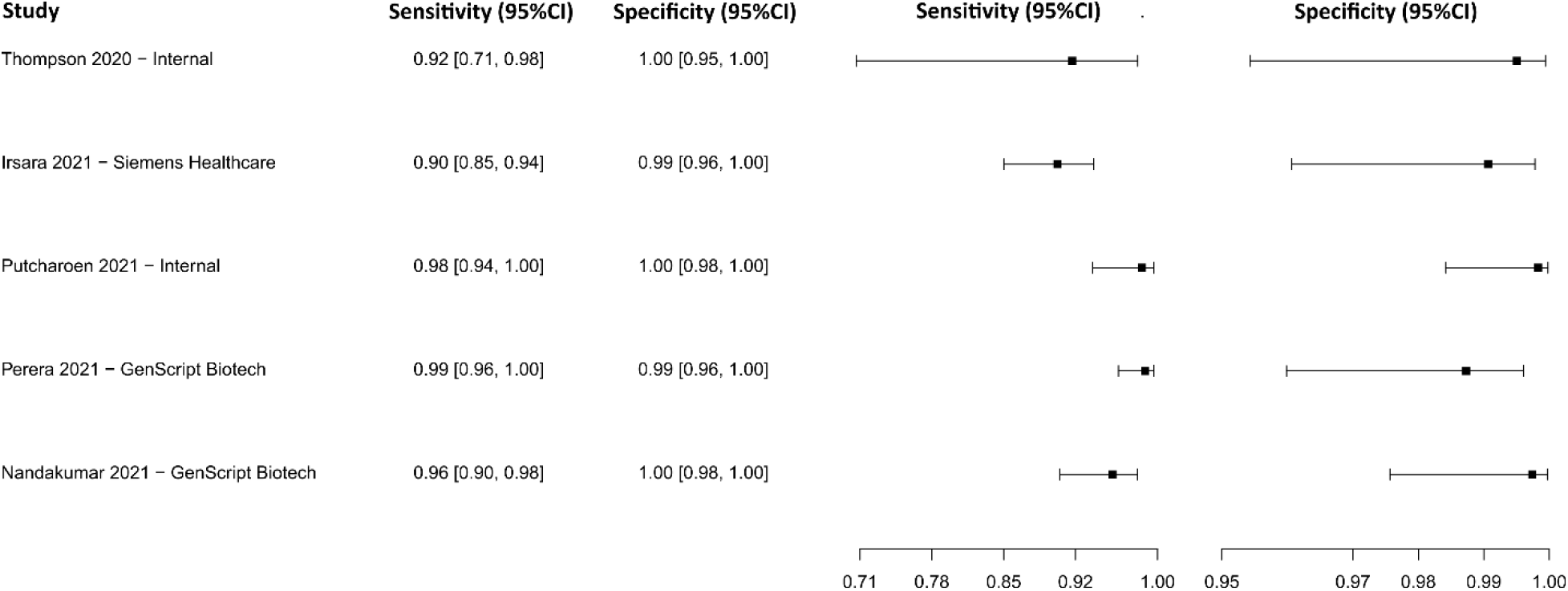
Data analysis and paired forest plot of the sensitivity and specificity of the antiviral neutralization bioassay (ANB) in the diagnosis of COVID-19. Sensitivity and specificity are reported as a mean (95% confidence limits). The forest plot represents the estimated sensitivity and specificity (black squares) and their 95% confidence limits (horizontal black line) [91–95].

#### Biosensors

Seven studies were selected using BS as a diagnostic technique [96–102]. A total of 814 subjects were studied. Sensitivity ranged from 90.0 to 98.8%, with a median of 96.4%, and CI of 95% (85.9, 99.2), while the test for equality of sensitivities presented a χ^2^ = 6.63, df = 6, p-value = 0.35. Specificity ranged from 89.3 to 99.5%, with a median of 97.4%, 95%CI (93.1, 99.5); the test for equality of specificities showed χ^2^ = 17.31, df = 6, p-value = 0.01. A negative correlation between sensitivities and false positive rates is shown r = 0.385, 95%CI (–0.882, 0.518). In addition, results regarding LR+ {median 36.15, 95%CI (8.24, 297.95)}, LR− {median 0.04, 95%CI (0.01, 0.19)} and DOR {median 459.96, 95%CI (129.02, 4295.06)}. The analyzed diagnostic performance is summarized in Figure 11 and Supplementary Figure 8.

**Figure 11.**
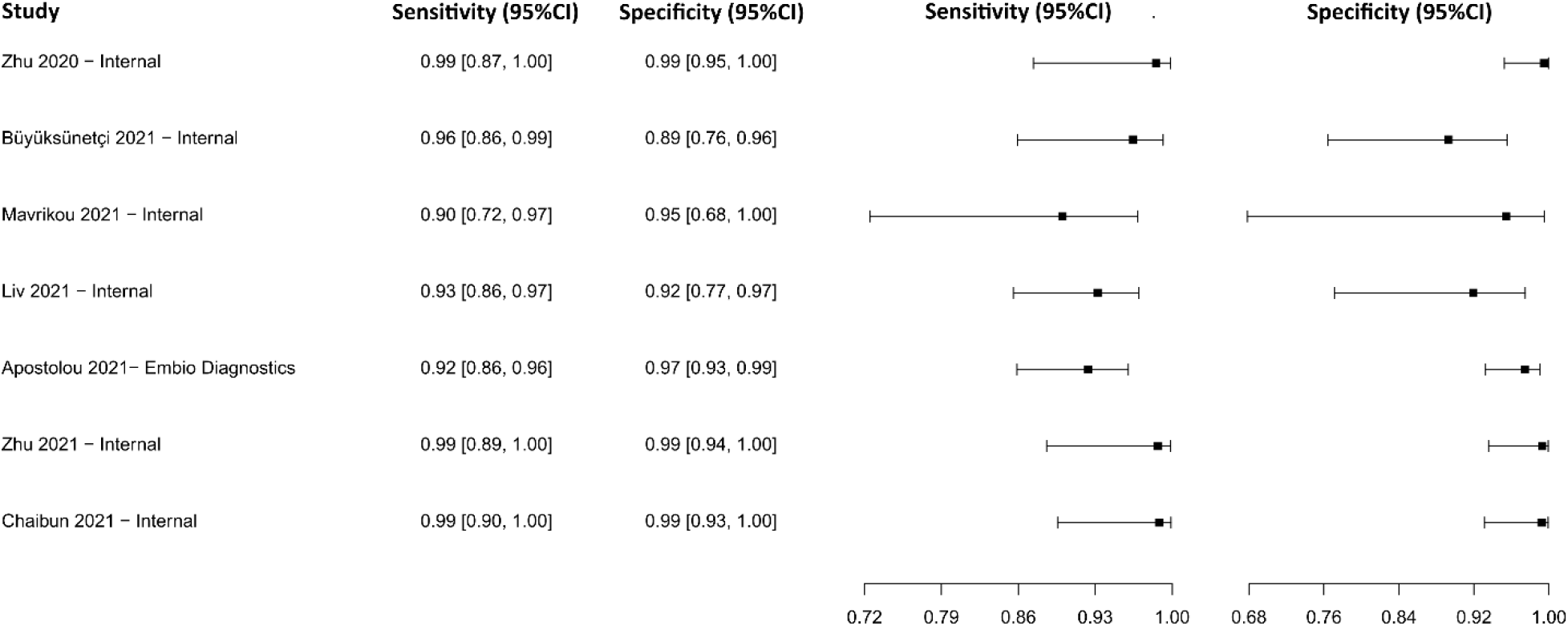
Data analysis and paired forest plot of the sensitivity and specificity of the biosensors (BS) in the diagnosis of COVID-19. Sensitivity and specificity are reported as a mean (95% confidence limits). The forest plot represents the estimated sensitivity and specificity (black squares) and their 95% confidence limits (horizontal black line) [96–102].

#### Chemiluminescence immunoassay for IgG

Eight studies were selected using CLIA as an IgG detection technique [76,77,79,88,103–107], in which a total of 2,859 subjects were studied. Sensitivity ranged from 53.1% to 96.5%, with a median of 79.8%, 95%CI (65.3, 89.7), while the test for equality of sensitivities presented a χ^2^ = 111.96, df = 11, p-value = 2e^-16^. Specificity ranged from 89.8 to 99.9%, with a median of 98.7%, 95%CI (93.5, 99.6); the test for equality of specificities showed χ^2^ = 47.84, df = 11, p-value = 1.53e^-06^. A negative correlation between sensitivities and false positive rates is shown r = 0.319, 95%CI (–0.312, 0.755). Additionally, results regarding LR+ {median 62.27, 95%CI (7.99, 248.31)}, LR− {median 0.21, 95%CI (0.11, 0.38)} and DOR {median 286.73, 95%CI (46.74, 2894.79)}. The analyzed diagnostic performance is summarized in Figure 12 and Supplementary Figure 9.

**Figure 12.**
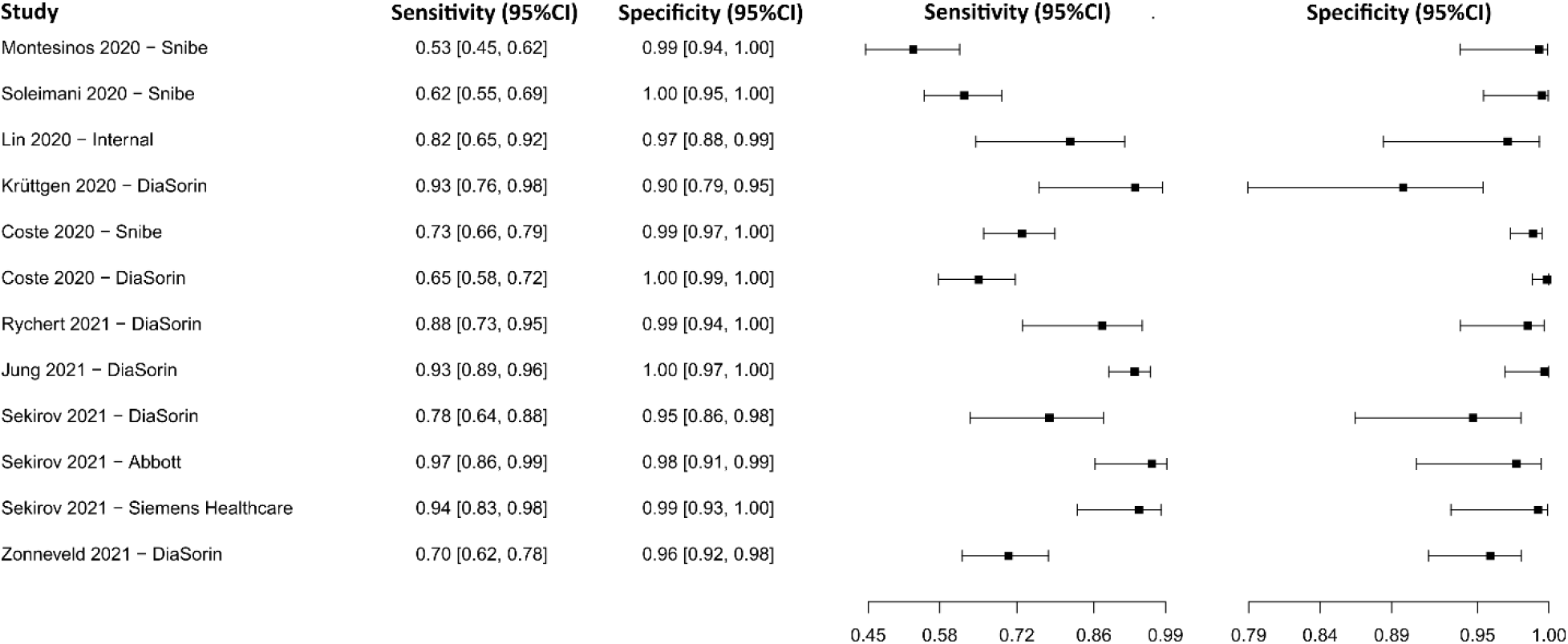
Data analysis and paired forest plot of the sensitivity and specificity of the chemiluminescence immunoassay (CLIA) for IgG in the diagnosis of COVID-19. Sensitivity and specificity are reported as a mean (95% confidence limits). The forest plot represents the estimated sensitivity and specificity (black squares) and their 95% confidence limits (horizontal black line) [76,77,79,88,103–107].

#### Chemiluminescence immunoassay for IgM

Five studies were selected using CLIA as an IgM detection technique [77,79,103,104,106]. A total of 1,240 subjects were studied. Sensitivity ranged from 58.7 to 89.5%, with a median of 61.7%, 95%CI (53.0, 77.0);), while the test for equality of sensitivities showed: χ^2^ = 18.52, df =4, p-value = 9.76e^-04^. Specificity ranged from 91.3 to 99.5%, with a median of 99.2%, 95%CI (94.0, 100.0); while the test for equality of specificities presented χ^2^ = 13.50, df = 4, p-value = 9.06e^-03^. The correlation between sensitivities and false positive rates was analyzed a negative result is shown r = -0.252, 95%CI (-0.928, 0.810). In addition, results regarding LR+ {median 85.65, 95%CI (7.46, 1361.54)}, LR− {median 0.40, 95%CI (0.27, 0.48)} and DOR {median 250.76, 95%CI (18.56, 3396.17)} are displayed. The analyzed diagnostic performances are summarized in Figure 13 and Supplementary Figure 10.

**Figure 13.**
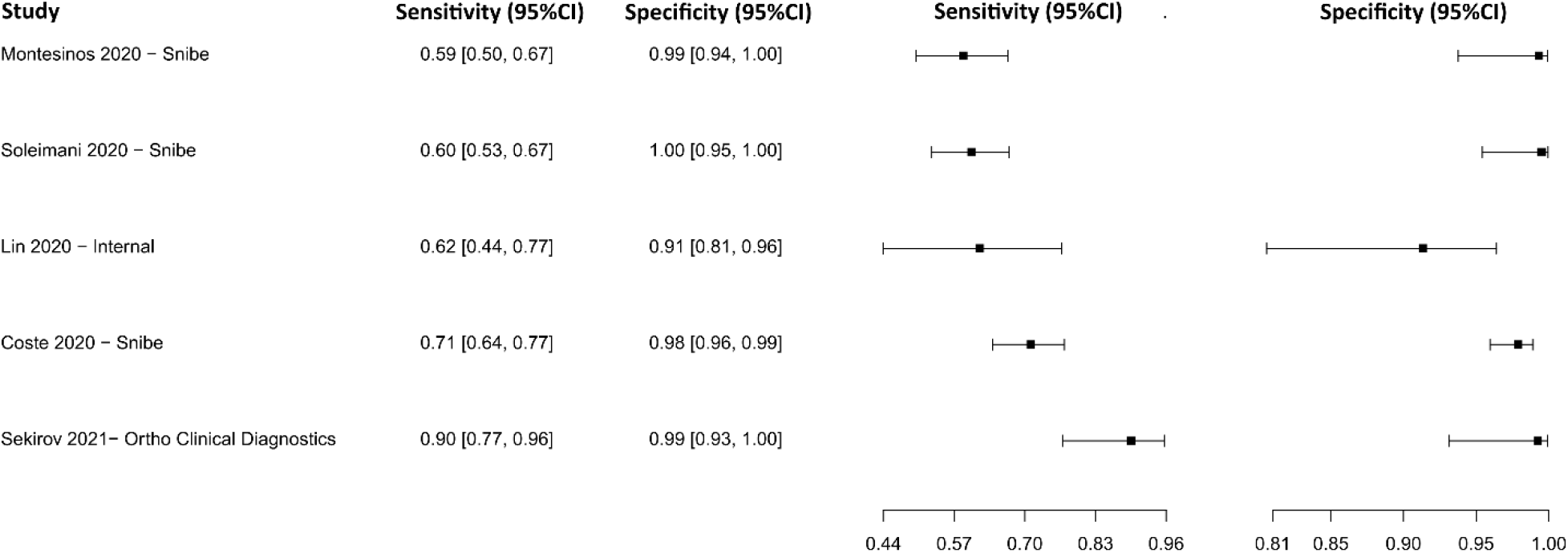
Data analysis and paired forest plot of the sensitivity and specificity of the chemiluminescence immunoassay (CLIA) for IgM in the diagnosis of COVID-19. Sensitivity and specificity are reported as a mean (95% confidence limits). The forest plot represents the estimated sensitivity and specificity (black squares) and their 95% confidence limits (horizontal black line) [77,79,103,104,106].

#### Chemiluminescence immunoassay for IgM-IgG

The analysis identified 28 published studies that used CLIA to detect IgM-IgG antibodies for COVID-19. After analysis, only five studies [77,85,103,106,108] were selected. A total of 1,008 subjects were studied. Sensitivity ranged from 64.2 to 98.8%, with a median of 90.1%, and 95%CI (80.0, 95.0). Test for equality of sensitivities analysis showed: χ^2^ = 51.25, df = 6, p-value = 2.64e^-09^. The specificity of the studies ranged from 97.7 to 99.5%, with a median of 99.2%, 95%CI (93.0, 100.0);), while the test for equality of specificities: χ^2^ = 1.73, df = 6, p-value = 0.94. Also, a negative correlation between sensitivities and false positive rates is shown r = 0.635, 95%CI (–0.226, 0.939). In addition, results regarding LR+ {median 106.45, 95%CI (7.03, 1685.99)}, LR− {median 0.10, 95%CI (0.05, 0.22)} and DOR {median 828.39, 95%CI (45.47, 15090.85)} are displayed. The diagnostic performance of the selected studies is summarized in Figure 14 and Supplementary Figure 11.

**Figure 14.**
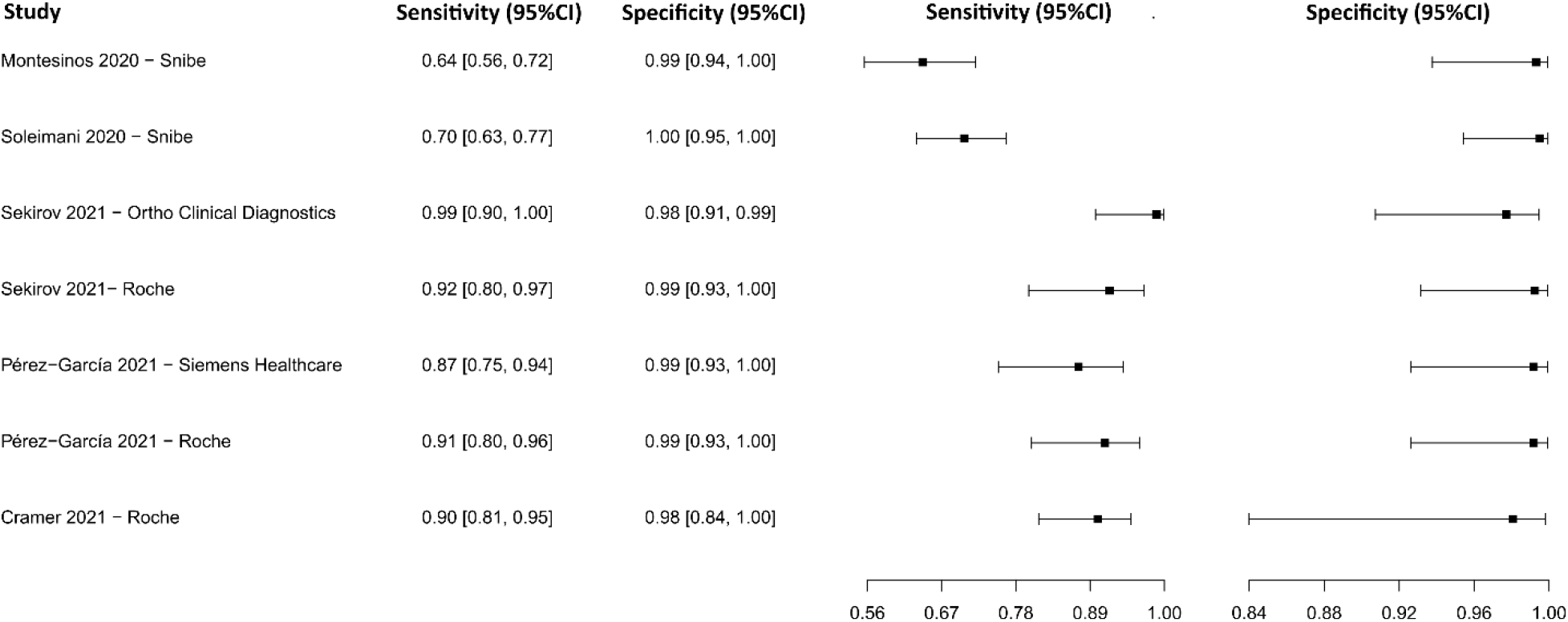
Data analysis and paired forest plot of the sensitivity and specificity of the chemiluminescence immunoassay (CLIA) for IgM-IgG in the diagnosis of COVID-19. Sensitivity and specificity are reported as a mean (95% confidence limits). The forest plot represents the estimated sensitivity and specificity (black squares) and their 95% confidence limits (horizontal black line) [77,85,103,106,108].

#### Lateral flow immunoassay for IgG

Eleven studies were selected using LFIA as an IgG detection technique [75,77,79,109–116]. A total of 15,935 subjects were studied. Sensitivity ranged from 35.9 to 97.4%, with a median of 87.3%, 95%CI (76.5, 91.9), while the test for equality of sensitivities presented a χ^2^ = 666.12, df = 21, p-value = 2e^-16^. Specificity ranged from 88.5 to 99.6%, with a median of 97.9%, 95%CI (95.5, 99.3); the test for equality of specificities showed χ^2^ = 48.01, df = 21, p-value = 6.85e^-04^. A negative correlation between sensitivities and false positive rates is shown r = 0.175, 95%CI (–0.267, 0.555). In addition, results regarding LR+ {median 40.07, 95%CI (11.54, 131.19)}, LR− {median 0.13, 95%CI (0.08, 0.25)} and DOR {median 502.97, 95%CI (56.53, 1796.41)}. The analyzed diagnostic performance is summarized in Figure 15 and Supplementary Figure 12.

**Figure 15.**
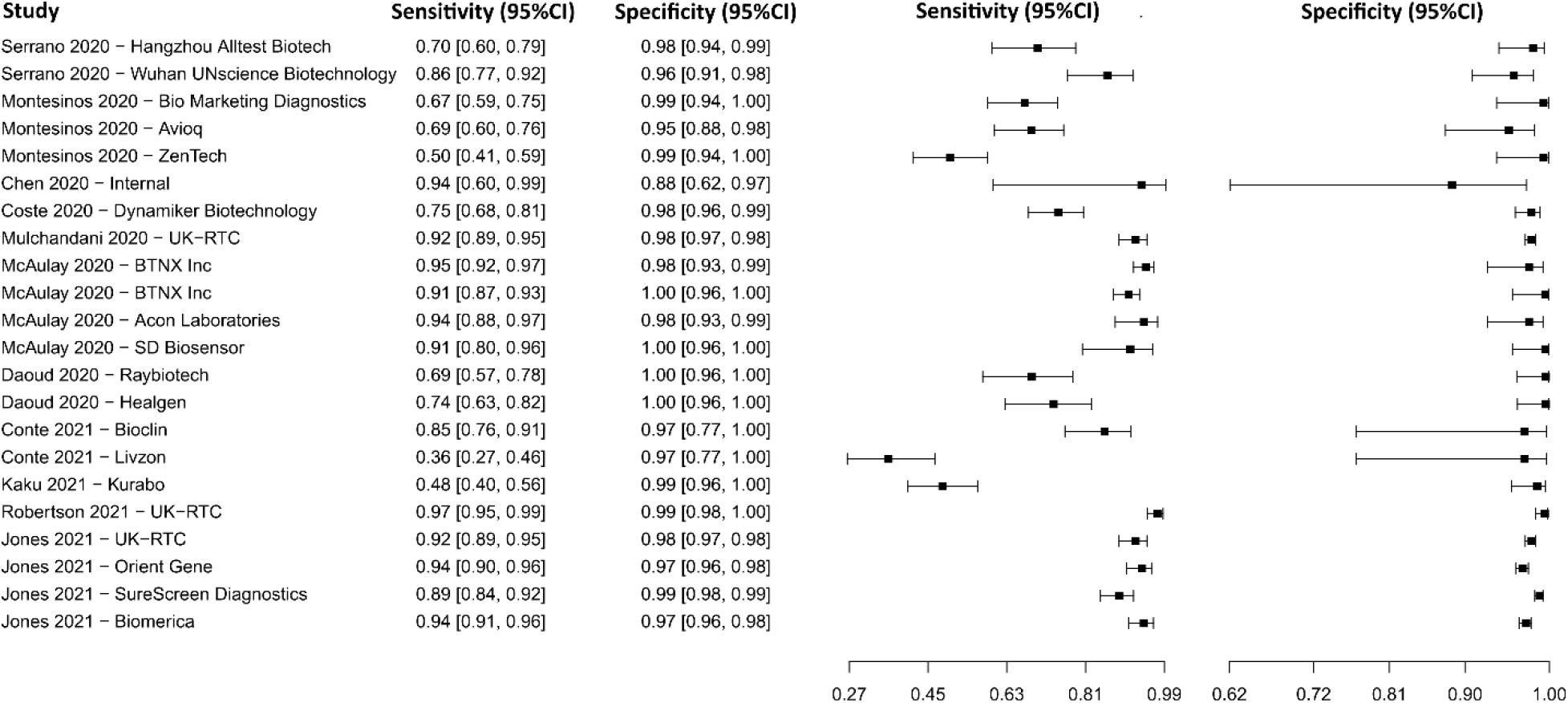
Data analysis and paired forest plot of the sensitivity and specificity of the lateral flow immunoassay (LFIA) for IgG in the diagnosis of COVID-19. Sensitivity and specificity are reported as a mean (95% confidence limits). The forest plot represents the estimated sensitivity and specificity (black squares) and their 95% confidence limits (horizontal black line) [75,77,79,109–116].

#### Lateral flow immunoassay for IgM

Six studies were selected using LFIA as an IgM detection technique [75,77,79,111,112,115], in which a total of 2,704 subjects were studied. Sensitivity ranged from 23.3% to 87.2%, with a median of 62.4%, 95%CI (51.1, 72.5), while the test for equality of sensitivities presented a χ^2^ = 208.82, df = 11, p-value = 2e^-16^. Specificity ranged from 89.7 to 99.7%, with a median of 98.0%, 95%CI (93.7, 99.7); the test for equality of specificities showed χ^2^ = 40.45, df = 11, p-value = 3.00e^-05^. A negative correlation between sensitivities and false positive rates is shown r = 0.153, 95%CI (–0.461, 0.669). Additionally, results regarding LR+ {median 33.70, 95%CI (5.07, 263.29)}, LR− {median 0.38, 95%CI (0.29, 0.51)} and DOR {median 96.36, 95%CI (13.50, 651.54)}. The analyzed diagnostic performance is summarized in Fig. 16 and Supplementary Figure 13.

**Figure 16.**
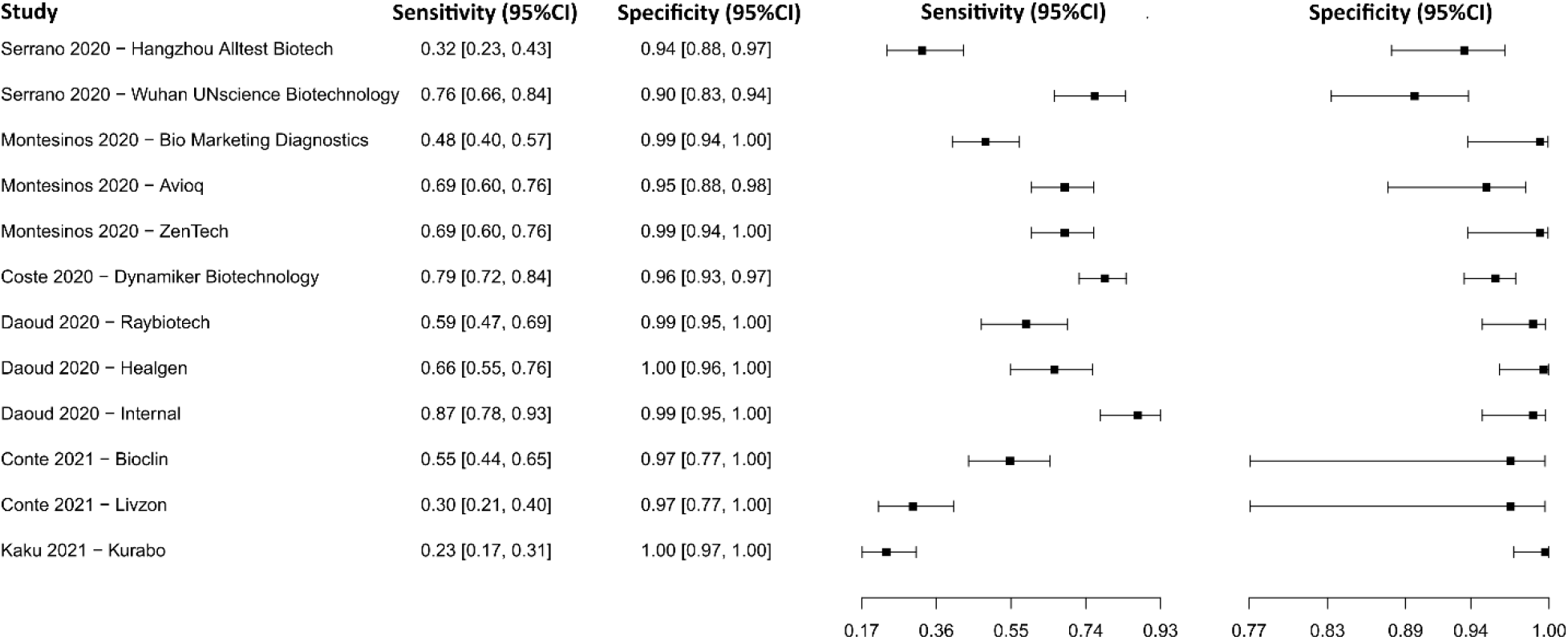
Data analysis and paired forest plot of the sensitivity and specificity of the lateral flow immunoassay (LFIA) for IgM in the diagnosis of COVID-19. Sensitivity and specificity are reported as a mean (95% confidence limits). The forest plot represents the estimated sensitivity and specificity (black squares) and their 95% confidence limits (horizontal black line) [75,77,79,111,112,115].

#### Lateral flow immunoassay for IgM-IgG

Nine studies were selected using LFIA as an IgM-IgG detection technique [75,77,85,90,108,109,115,117,118]. A total of 9,629 subjects were studied. Sensitivity ranged from 44.1 to 97.0%, with a median of 83.7%, 95%CI (63.4, 88.2);), while the test for equality of sensitivities showed: χ^2^ = 339.59, df =20, p-value = 2e^-16^. Specificity of the studies ranged from 87.4 to 99.5%, with a median of 97.1%, 95%CI (92.4, 99.7); while the test for equality of specificities presented χ^2^ = 107.85, df = 20, p-value = 4.83e^-14^. The correlation between sensitivities and false positive rates was analyzed a negative result is shown r = 0.279, 95%CI (-0.173, 0.635). In addition, results regarding LR+ {median 30.97, 95%CI (6.32, 445.62)}, LR− {median 0.18, 95%CI (0.13, 0.39)} and DOR {median 334.86, 95%CI (22.18, 2704.72)} are displayed. The analyzed diagnostic performances are summarized in Figure 17 and Supplementary Figure 14.

**Figure 17.**
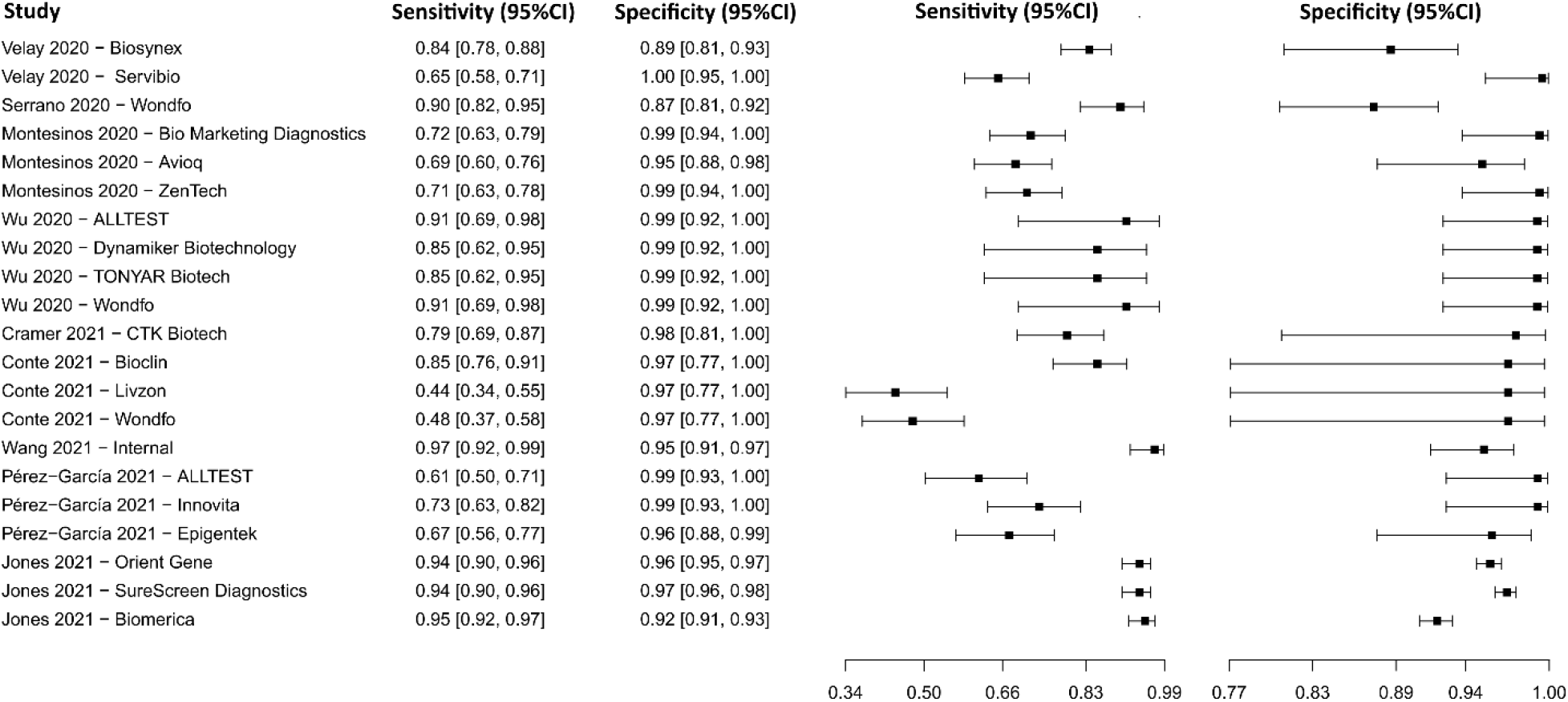
Data analysis and paired forest plot of the sensitivity and specificity of the lateral flow immunoassay (LFIA) for IgM-IgG in the diagnosis of COVID-19. Sensitivity and specificity are reported as a mean (95% confidence limits). The forest plot represents the estimated sensitivity and specificity (black squares) and their 95% confidence limits (horizontal black line) [75,77,85,90,108,109,115,117,118].

#### Lateral flow immunoassay for N protein

Fourteen studies were selected using LFIA as an N protein detection technique [119–132]. A total of 11,750 subjects were studied. Sensitivity ranged from 18.3 to 96.9%, with a median of 74.7%, 95%CI (50.7, 88.3), while the test for equality of sensitivities presented a χ^2^ = 145.62, df = 21, p-value = 2e^-16^. Specificity ranged from 93.8 to 99.9%, with a median of 99.4%, 95%CI (95.9, 99.8); the test for equality of specificities showed χ^2^ = 56.33, df = 21, p-value = 4.51e^-05^. A negative correlation between sensitivities and false positive rates is shown r = 0.193, 95%CI (–0.249, 0.568). In addition, results regarding LR+ {median 85.90, 95%CI (12.25, 482.92)}, LR− {median 0.26, 95%CI (0.12, 0.54)} and DOR {median 501.51, 95%CI (46.03, 2611.02)}. The analyzed diagnostic performance is summarized in Figure 18 and Supplementary Figure 15.

**Figure 18.**
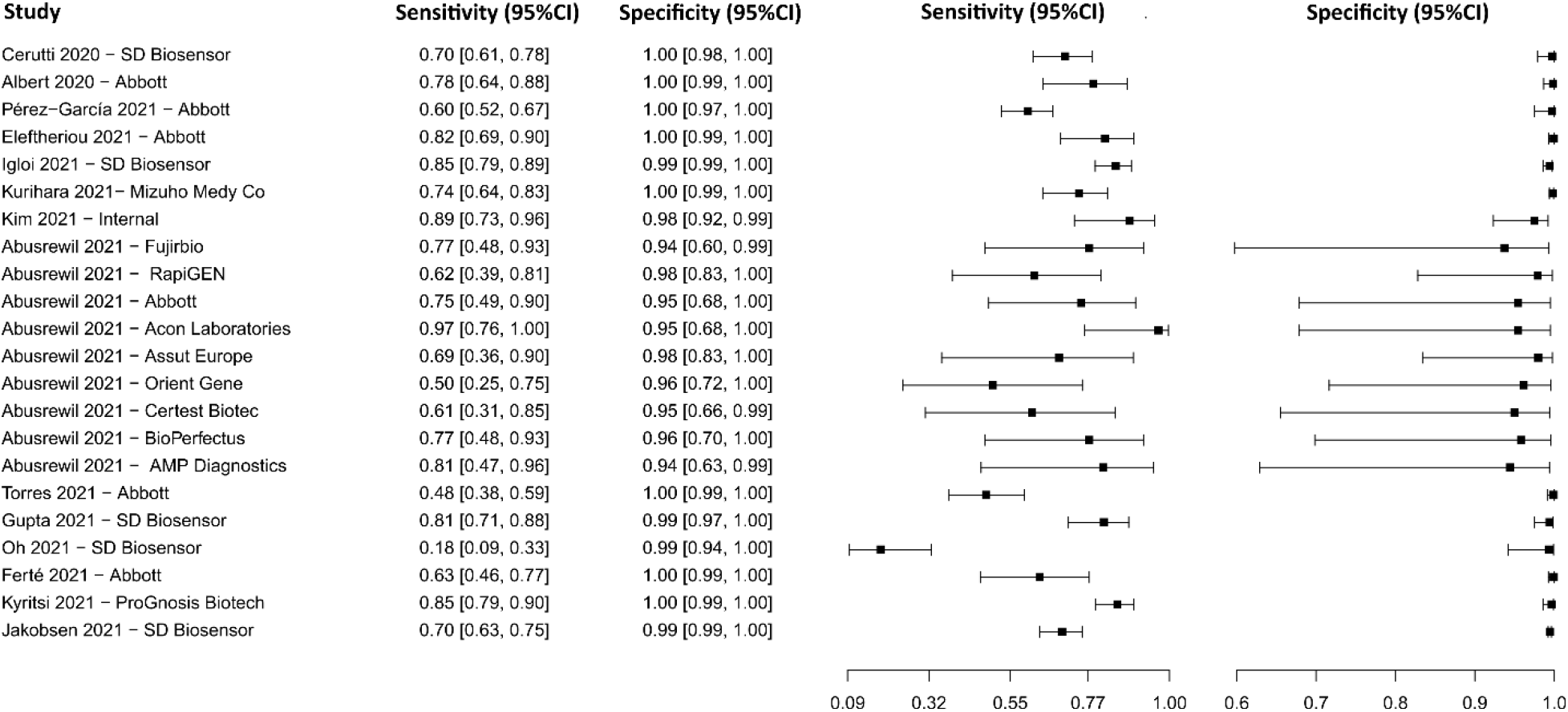
Data analysis and paired forest plot of the sensitivity and specificity of the lateral flow immunoassay (LFIA) for N-protein in the diagnosis of COVID-19. Sensitivity and specificity are reported as a mean (95% confidence limits). The forest plot represents the estimated sensitivity and specificity (black squares) and their 95% confidence limits (horizontal black line) [119–132].

#### Chemiluminescent microparticle immunoassay

The analysis identified 13 published studies that used CMIA as a diagnostic technique for COVID-19. After analysis, only five studies [81,88,105,107,133] were selected. A total of 939 subjects were studied. Sensitivity ranged from 62.8 to 95.7%, with a median of 90.3%, and 95%CI (76.4, 96.4). Test for equality of sensitivities analysis showed: χ^2^ = 51.58, df = 4, p-value = 1.69e^-10^. Specificity ranged from 95.3 to 99.7%, with a median of 98.8%, 95%CI (93.8, 99.8);), while the test for equality of specificities: χ^2^ = 6.26, df = 4, p-value = 0.18. Also, a negative correlation between sensitivities and false positive rates is shown r = 0.487, 95%CI (–0.693, 0.958). In addition, results regarding LR+ {median 60.79, 95%CI (12.37, 476.78)}, LR− {median 0.10, 95%CI (0.04, 0.27)} and DOR {median 615.95, 95%CI (74.57, 4343.60)} are displayed. The diagnostic performance of the selected studies is summarized in Figure 19 and Supplementary Figure 16.

**Figure 19.**
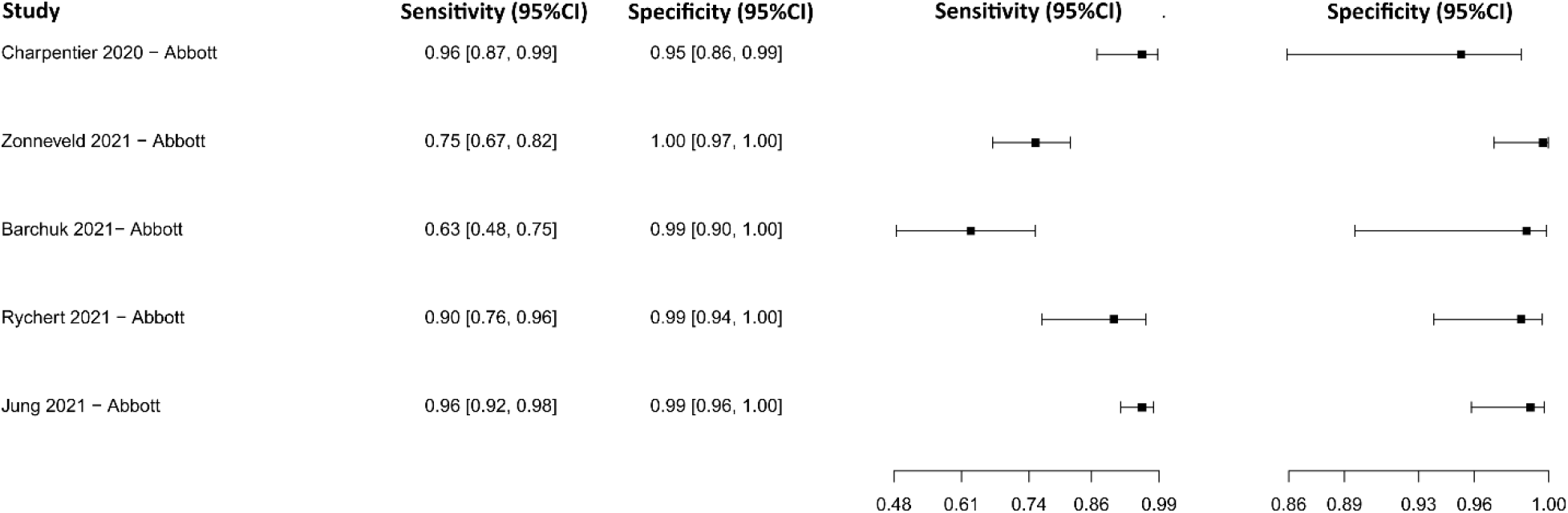
Data analysis and paired forest plot of the sensitivity and specificity of the chemiluminescent microparticle immunoassay (CMIA) in the diagnosis of COVID-19. Sensitivity and specificity are reported as a mean (95% confidence limits). The forest plot represents the estimated sensitivity and specificity (black squares) and their 95% confidence limits (horizontal black line) [81,88,105,107,133].

#### Fluorescence immunoassay

Three studies were selected using the FIA technique [121,134,135], in which a total of 829 subjects were studied. Sensitivity ranged from 38.0% to 92.6%, with a median of 64.4%, 95%CI (59.0, 73.0), while the test for equality of sensitivities presented a χ^2^ = 49.92, df = 4, p-value = 3.75e^-10^. Specificity ranged from 97.1 to 99.5%, with a median of 99.0%, 95%CI (93.5, 99.9); the test for equality of specificities showed χ^2^ = 4.12, df = 4, p-value = 0.39. A negative correlation between sensitivities and false positive rates is shown r = 0.282, 95%CI (–0.799, 0.932). Additionally, results regarding LR+ {median 77.43, 95%CI (5.56, 1275.84)}, LR− {median 0.35, 95%CI (0.28, 0.43)} and DOR {median 124.95, 95%CI (26.33, 2224.82)}. The analyzed diagnostic performance is summarized in Figure 20 and Supplementary Figure 17.

**Figure 20.**
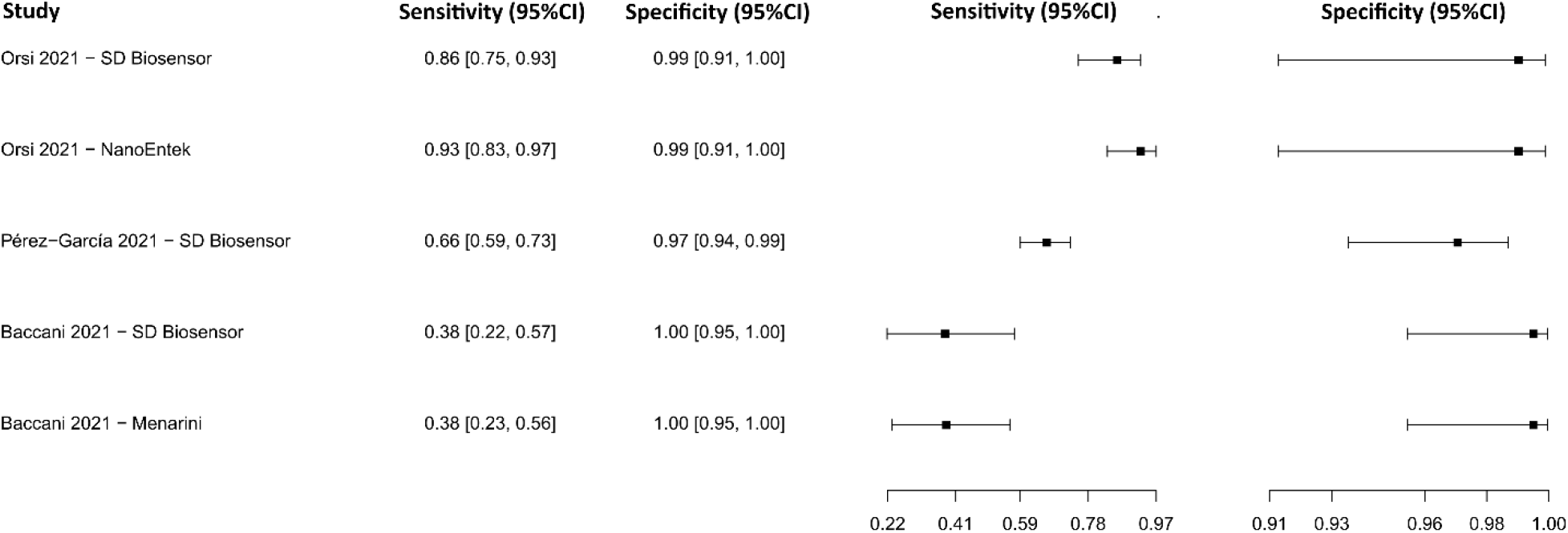
Data analysis and paired forest plot of the sensitivity and specificity of the fluorescence immunoassay (FIA) in the diagnosis of COVID-19. Sensitivity and specificity are reported as a mean (95% confidence limits). The forest plot represents the estimated sensitivity and specificity (black squares) and their 95% confidence limits (horizontal black line) [121,134,135].

#### Other techniques

Regarding the NGS, MA, ELISA for IgG-IgM-IgA, ELISA for IgG-IgM / IgG-IgA, CLIA for IgG-IgM-IgA, CLIA for N protein, LFIA for S protein and ECLIA diagnostic techniques, one [89], zero, four [74,81,136,137], two [77,108], one [137], four [135,138–140], one [126], and three [105,107,111] studies were selected, respectively. According to the established criteria, at least five studies were needed for the analysis with a value of p < 0.05. So, no analysis was developed regarding these diagnostic techniques.

#### Summary ROC curves (sROC)

Comparison of the diagnostic techniques data for COVID-19 (RT-PCR, RT-LAMP, CRISPR, ELISA IgG, ELISA IgM, ELISA IgA, ABN, BS, CLIA IgG, CLIA IgM, CLIA IgM-IgG, LFIA IgG, LFIA IgM, LFIA IgM-IgG, LFIA N protein, CMIA, and FIA) was performed through an sROC curve analysis (Figure 21), due to implicit or explicit alterations between studies and variation in the cut-off points of the test, differences in sensitivity and specificity may occur [141,142]. The area under the curve (AUC) calculated for the diagnostic techniques for COVID-19 is shown in Figure 21, showing better performance for ABN. Furthermore, when the AUC was limited to the observed false positive rates (FPR) (AUC_FPR_) results revealed the relatively better performance of the ABN diagnostic test for COVID-19 (Figure 21).

**Figure 21.**
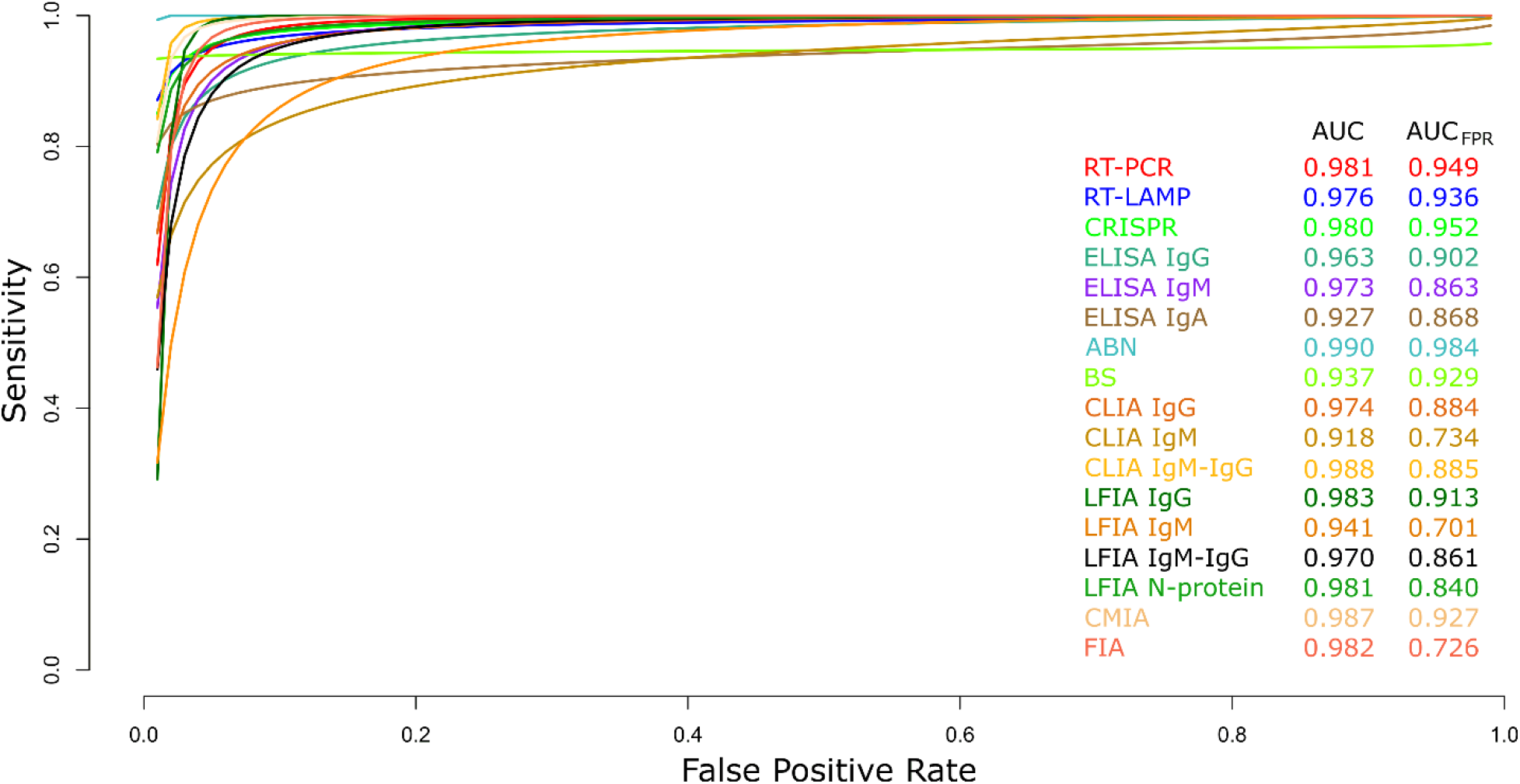
Meta-analysis of diagnostic test accuracy analysis. Summary receiver operating curve (sROC) plot of false positive rate and sensitivity. Comparison between RT-PCR, RT-LAMP, CRISPR, ELISA IgG, ELISA IgM, ELISA IgA, ABN, BS, CLIA IgG, CLIA IgM, CLIA IgM-IgG, LFIA IgG, LFIA IgM, LFIA IgM-IgG, LFIA N protein, CMIA and FIA methods in the diagnosis of COVID-19.

## Discussion

COVID-19 is a disease that has caused a devastating impact, leaving an experience for the new generations to take into consideration different factors that favor the emergence of infectious diseases [143]. The COVID-19 pandemic highlighted the weaknesses that exist in disease detection, alert, and response mechanisms. It demonstrated the need for a restart of the global health and health security system, as the world is highly interconnected, and a pandemic can break out in a matter of days [144]. The transmission of infectious diseases is mainly related to human movement, in the increase is related to transportation networks and globalization. In this context, the spread of pathogenic microorganisms, such as SARS-CoV-2, can be rapid and difficult its prevent and manage [145]. Furthermore, the ratio of asymptomatic people who give a positive result for COVID-19 varies between 8.44% and 39.00% [146], with the potential that asymptomatic carriers could be capable of transmitting COVID-19 during the incubation process without showing any symptoms or sign [147,148]. The viral load of SARS-CoV-2 decreases rapidly with the immune response; on the other hand, the generation of antibodies begins days after the onset of the infection, so to establish an adequate diagnosis, the start time of the possible infection should be considered. infection, taking into account that the sensitivity of molecular tests will decrease over time, but the sensitivity of serological tests will increase after several days of infection[149]. For these reasons, the accurate and timely identification of patients infected with COVID-19 plays an important role in the detection, diagnosis, follow-up, and surveillance of this pandemic [150,151]. However, most of the early diagnostic techniques developed for COVID-19 present limitations, such as sensitivity and/or specificity [152]. In addition, diagnostic tests required processing in laboratories with sophisticated materials, leading to a longer turnaround time and several days needed for results be available. Some of these hurdles were later perfected by newly developed assays that have better-defined analytical accuracy than earlier assays with better-defined analytical accuracy than earlier one[153,154].

SARS-CoV-2 is an RNA virus and, therefore, the different molecular tests available for detection of R; therefore, the different molecular tests available for detecting can be used to identify the virus [155,156]. RNA of the virus must be transcribed into a DNA complement by reverse transcriptase to apply DNA detection methods [157]. Currently, the most widely used molecular method for the early detection of COVID-19 is RT-PCR; while other promising options, such as RT-LAMP and CRISPR have been developed [158]. RT-PCR is considered the gold standard molecular diagnostic test for COVID-19 worldwide due to its high specificity; however, limited access to kits and reagents, the use of expensive laboratory equipment, and the need for qualified personnel led to slow detection of the virus [159,160]. As well, it has been reported that the RT-PCR test can fail in suspected and confirmed patients with clinical implications. In this context, other clinical and molecular tests should also be considered in the diagnosis of COVID-19 [159,161]. On the other hand, it has been described that asymptomatic infected people have a few copies below the detection limit of the nucleic acid within the upper respiratory tract, causing false negatives in asymptomatic infected individuals, due to this the determination of asymptomatic infections with high specificity and sensitivity is a key point for managing the pandemic [162]. About these criteria, the analysis showed that there is no significant difference between molecular tests between RT-PCR and CRISPR, with a median sensitivity of 94.5, 94.4%, and specificity of 98.4, and 98.6% for RT-PCR. PCR, and CRISPR, respectively. However, when comparing AUC_FPR_, CRISPR showed superior results, which can be explained partially explained by the difference in the number of studies evaluated and sample sizes. CRISPR-based COVID-19 diagnosis has advantages such as high detection speed (approximately 30 minutes), high sensitivity and accuracy, portability, and does not need specialized laboratory equipment [160]. Furthermore, RT-LAMP showed the highest specificity among the molecular tests with a median of 98.8% but the lowest sensitivity with a median of 91.9%. RT-LAMP has advantages over RT-PCR, such as amplification at a constant temperature, no thermocycler required, and a faster result, while maintaining similar sensitivity and specificity, which could make it more suitable than RT-PCR for monitoring a pandemic [163]. The molecular diagnosis makes it possible to identify people infected with SARS-CoV-2 one week before the onset of symptoms, while. At the same time, antibodies can only be detected ≥ 8 days after the onset of symptoms, so molecular diagnoses are essential for early diagnosis of COVID-19 [164,165]. When comparing data from molecular tests with serological tests for the diagnosis of COVID-19, molecular tests in general present a better performance, with AUC_FPR_ of 0.949, 0.936, 0.952 for RT-PCR, RT-LAMP, and CRISPR, respectively.

There are various serological tests to detect the antibody response caused by COVID-19. The main methods are ELISA, CLIA, LFIA, CMIA, FIA, and ANB [166–168]. It has been reported that serological assays have adequate sensitivity and specificity for the detection of SARS-CoV2, but this performance is related to the patient’s convalescence, so understanding the kinetics of antibodies during SARS-CoV-2 infection. In this context, it is essential to establish the serological result for the diagnosis of the disease [168,169]. Data from ELISA, CLIA, LFIA, CMIA, and FIA-based serological tests show low sensitivity with a median less than or equal to 90% and high specificity with a median greater than or equal to 97%. Low sensitivity would produce false negative results since they cannot detect intrinsic immunological differences or immune responses between individuals, and it must be considered that in asymptomatic patients antibodies can be produced, still, their titers are not as high as those detected in symptomatic patients [170]. When SARS-CoV-2 invades the human body and releases antigens, the immune system is activated to produce many specific antibodies (IgM/IgA/IgG), the host’s immune cells produce IgM and IgA at an early stage of infection, while IgG at a late stage [171–173].

ELISA, CLIA, and LFIA diagnostic techniques were broken down according to the antibody they detected (IgG, IgM, IgA, and IgG-IgM). The detection of IgG by these tests generated the highest AUC_FPR_ demonstrating the higher performance of this antibody. This could be because IgM levels start to decrease at week 5 and almost disappear after week 7, while IgG levels persist beyond week 7, demonstrating the greater stability of IgG [174]. On the other hand, different structural proteins are anchored on the surface of the SARS-CoV-2 membrane, mainly spike (S) nucleocapsid (N), membrane (M), and envelope (E) [173,175]; N protein is a highly immunogenic protein and abundantly expressed during infection, so it can be useful for the diagnosis of COVID-19 [176]. In the data analysis of the LFIA technique, its performance was analyzed against the detection of detecting the N protein of SARS-COV-2, giving a median sensitivity and specificity of 74.7% and 99.4%, respectively. Besides, it should be noted that N protein has better stability than RNA, which can effectively compensate for the low sensitivity it shows [177]. Because many companies can generate serology tests, these tests can reach millions of people per day, which can help improve the detection of SARS-CoV-2, especially in countries with limited resources [177]. However, to obtain adequate sensitivity and specificity, it is recommended to combine clinical, molecular, and serological diagnostic tests [178].

It is worth noting that the diagnostic test that showed a higher performance for the detection of COVID-19, considering molecular and serological techniques, was the ANB, with a median of 95.6 and 99.5% for sensitivity and specificity, respectively; in addition, it obtained the highest AUC_FPR_ of 0.984. Neutralizing antibodies are produced weeks after infection and can protect cells from virus intrusion, protecting cells from virus intrusion and giving cells protective immunity [179]. ANBs are capable of quantitatively detecting neutralizing antibodies against SARS-CoV-2, making it possible to analyze the relationship between the level of neutralizing antibodies and the severity of the disease, and they can also predict the possibility of reinfection in patients with COVID-19 [179,180]. In general, neutralizing antibodies are measured by the plaque reduction neutralization test (PRNT), which is currently the gold standard for serological tests and the determination of immune protection [181].

Molecular techniques such as NGS allow the investigation of the genetic composition of SARS-CoV-2, allowing its detection [182], and MA has been used for genotyping and the determination of agents that cause diseases such as SARS-CoV-2 [183,184]. Furthermore, ECLIA serological-based diagnostic technique is a valid detection method for COVID-19 [185,186]; however, the number of selected studies of these techniques prevented their inclusion in the meta-analysis, which needs at least five studies for analysis with a p-value lower than 0.05 [142]. In a search of individual MeSH terms for “COVID-19”, “Sensitivity and specificity”, “Next generation sequencing”, “Microarray Analysis”, and “Immunoassay” showed 195,931, 641,415, 49,966, 95,596, and 2,150 studies, respectively, while linking them only found 16, 3 and 367, correspondingly. For what should be considered as work limitations, common errors associated with systematic review and meta-analysis studies, such as study location and selection, missing important information about the results, inappropriate subgroup analysis, conflicts with new experimental data, and duplicate publication [187]. In addition, various problems, such as heterogeneity of study groups, clinical settings, and measures of diagnostic performance, were established in the present study, while. In contrast, biased estimates of diagnostic test performance could be overestimated [142,188]. It should be considered that to obtain adequate sensitivity and specificity, it is recommended to combine clinical, molecular, and serological diagnostic tests [178]; however, applying all these diagnostic techniques simultaneously is a barrier for countries with populations living in poverty, clearly observing the inequity in health that exists, which is a problem for the control of any pandemic [189].

## Conclusions

The precise detection of new infectious agents such as SARS-CoV-2 is important for the proper management of properly managing a pandemic to stop its spread. The present study evaluated the performance of various diagnostic techniques reported for COVID-19, where molecular tests (RT-PCR, RT-LAMP, and CRISPR) presented a better performance in terms of sensitivity and specificity as compared to serological tests (ELISA, CLIA, LFIA, CMIA, and FIA) for the detection of SARS-CoV-2. In addition, serological tests were shown to have high specificity, mainly when IgG was detected, but relatively low sensitivity. It should be noted that the ANB-based diagnostic technique reported the best performance among all the methods evaluated, demonstrating its potential for the diagnosis of SARS-CoV-2 infection. However, further optimization of testing, in general, is still required, and emphasis should be placed on the development of rapid, scalable, and high-precision assays for the control of future outbreaks of SARS-CoV-2 and other infectious agents that may occur in the future.

## Data Availability

All data produced in the present study are available upon reasonable request to the authors

## Author Contributions

Conceptualization: M.A.C.-P. and M.A.C.-F.; data curation: M.A.C.-P., J. J. V. A., K. C. M., and L. D. G. M.; formal analysis: M.A.C.-P. and M.A.C.- F.; funding acquisition: M.A.C.-P. and M.A.C.-F.; investigation: L. D. G. M., A.S.G., R.C.G., R. A. M.D., and E.A.F.C.; methodology: M.A.C.-P. and M.A.C.-F.; writing—review and editing: A.S.G., R.C.G., R. A. M.D., E.A.F.C., and M.A.C.-F. All authors have read and agreed to the published version of the manuscript.

## Institutional Review Board Statement

Not applicable.

## Informed Consent Statement

Not applicable.

## Data Availability Statement

Not applicable.

## Acknowledgments

Not applicable.

## Abbreviations

The following abbreviations are used in this study.

ACE2: Angiotensin-converting enzyme 2
ANB: Antiviral neutralization bioassay
AUC: Area Under the Curve
AUC_FPR_: Area Under the Curve Restricted to The False Positive Rates
BS: Biosensors
CI: Confidence interval
CLIA: Chemiluminescence immunoassay
CMIA: Chemiluminescent microparticle immunoassay
COVID-19: Coronavirus disease
CRISPR: Clustered regularly interspaced short palindromic repeats
DOR: Diagnostic Likelihood Ratio
ECLIA: Electrochemiluminescence immunoassay
ELISA: Enzyme-Linked Immunosorbent Assay
FIA: Fluorescence immunoassay
FN: False negatives
FP: False positives
IgA: Immunoglobulin A
IgG: Immunoglobulin G
IgM: Immunoglobulin M
INPLASY: International Platform of Registered Systematic Review and Meta-analysis Protocols
LFIA: Lateral flow immunoassay
LR-: Negative Likelihood Ratio
LR+: Positive Likelihood Ratio
MA: Microarrays
MeSH: Medical Subject Headings
NCBI: National Center for Biotechnology Information
NGS: Next-generation sequencing
PRISMA: Preferred Reporting Items for Systematic Reviews and Meta-Analyses
ROC: Receiver Operating Characteristics
RT-LAMP: Reverse transcriptase loop-mediated isothermal amplification
RT-PCR: Reverse transcription–polymerase chain reaction
SARS-CoV-2: Severe acute respiratory syndrome coronavirus 2
Se: Sensibility
Sp: Specificity
sROC: Summary Receiver Operating Characteristics
TN: True negatives
TP: True positives
WHO: World Health Organization

## Figure Legends

**Supplementary 1.**
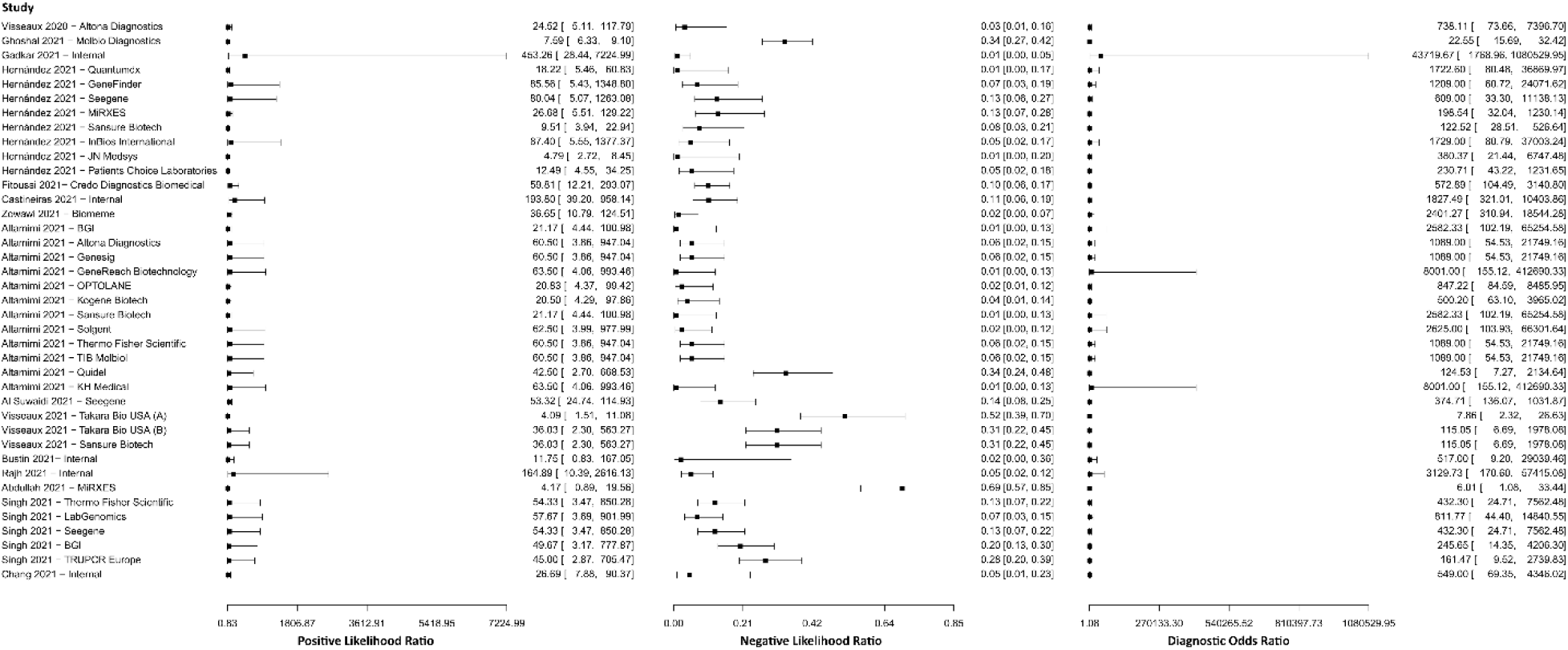
Study data and paired forest plot of the Positive Likelihood ratio, Negative likelihood ratio, and Diagnostic Odds ratio of reverse transcription–polymerase chain reaction (RT-PCR) in the diagnosis of COVID-19. Positive Likelihood ratio, Negative likelihood ratio, and Diagnostic Odds ratio are reported with a mean (95% confidence limits). The Forest plot depicts the estimated sensitivity and specificity (black squares) and its 95% confidence limits (horizontal black line) [44–58].

**Supplementary 2.**
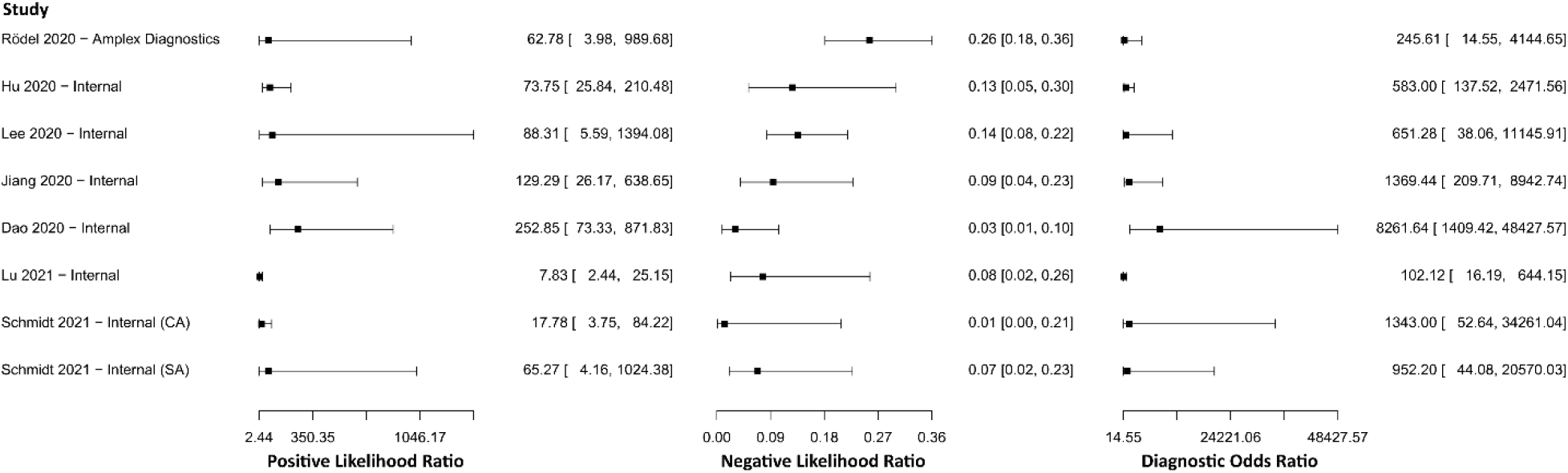
Study data and paired forest plot of the Positive Likelihood ratio, Negative likelihood ratio, and Diagnostic Odds ratio of reverse transcriptase loop-mediated isothermal amplification (RT-LAMP) in the diagnosis of COVID-19. Positive Likelihood ratio, Negative likelihood ratio, and Diagnostic Odds ratio are reported with a mean (95% confidence limits). The Forest plot depicts the estimated sensitivity and specificity (black squares) and its 95% confidence limits (horizontal black line) [59–65].

**Supplementary 3.**
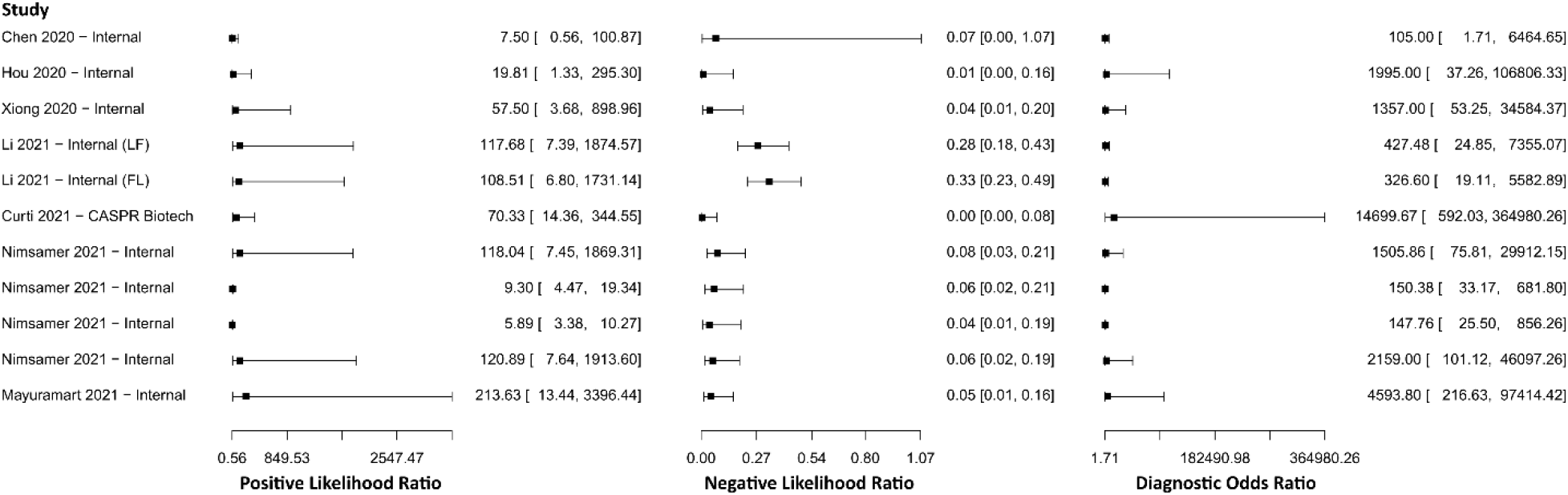
Study data and paired forest plot of the Positive Likelihood ratio, Negative likelihood ratio, and Diagnostic Odds ratio of clustered regularly interspaced short palindromic repeats (CRISPR) in the diagnosis of COVID-19. Positive Likelihood ratio, Negative likelihood ratio, and Diagnostic Odds ratio are reported with a mean (95% confidence limits). The Forest plot depicts the estimated sensitivity and specificity (black squares) and its 95% confidence limits (horizontal black line) [66–72].

**Supplementary 4.**
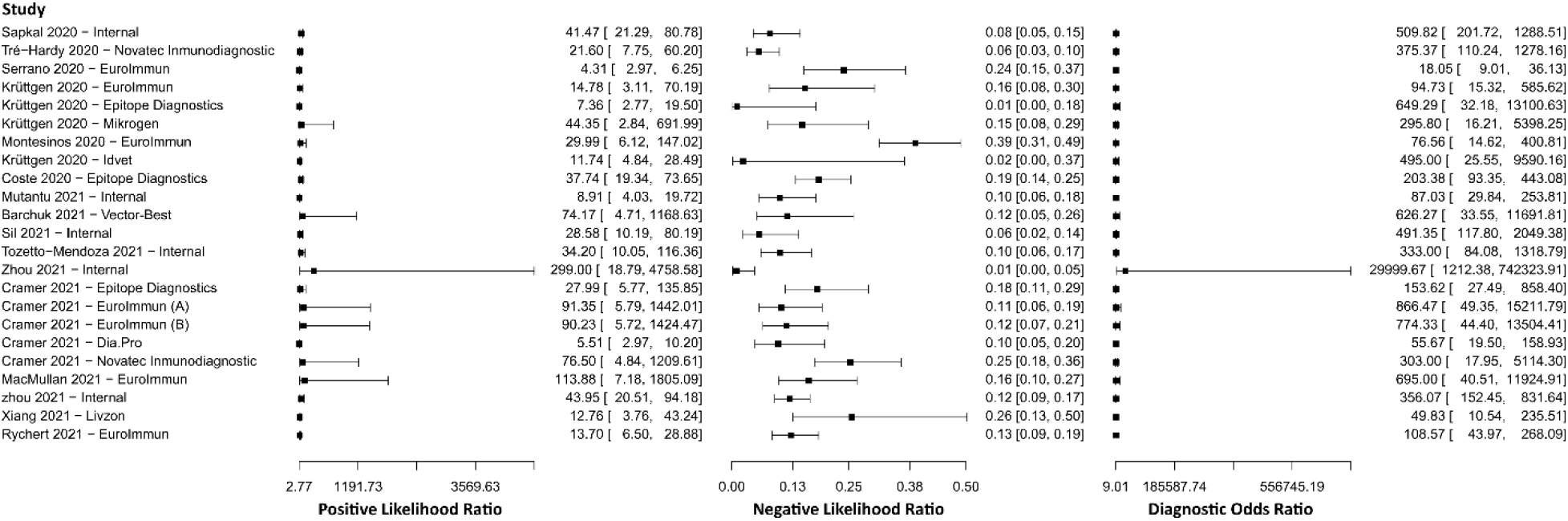
Study data and paired forest plot of the Positive Likelihood ratio, Negative likelihood ratio, and Diagnostic Odds ratio of enzyme-linked immunosorbent assay (ELISA) for IgG in the diagnosis of COVID-19. Positive Likelihood ratio, Negative likelihood ratio, and Diagnostic Odds ratio are reported with a mean (95% confidence limits). The Forest plot depicts the estimated sensitivity and specificity (black squares) and its 95% confidence limits (horizontal black line) [73–88].

**Supplementary 5.**
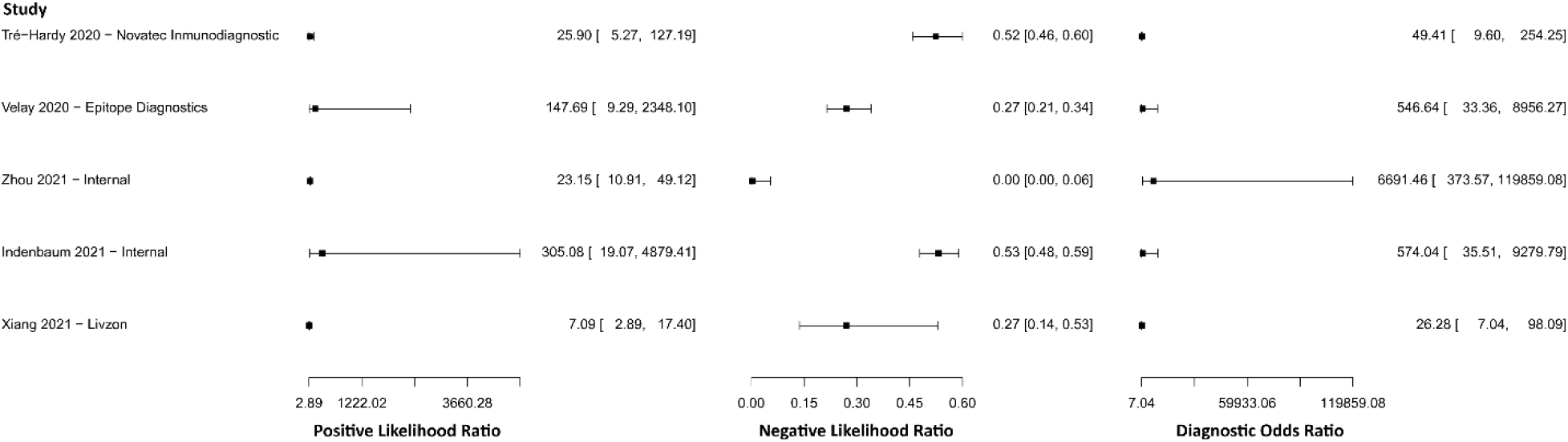
Study data and paired forest plot of the Positive Likelihood ratio, Negative likelihood ratio, and Diagnostic Odds ratio of enzyme-linked immunosorbent assay (ELISA) for IgM in the diagnosis of COVID-19. Positive Likelihood ratio, Negative likelihood ratio, and Diagnostic Odds ratio are reported with a mean (95% confidence limits). The Forest plot depicts the estimated sensitivity and specificity (black squares) and its 95% confidence limits (horizontal black line) [74,84,87,89,90].

**Supplementary 6.**
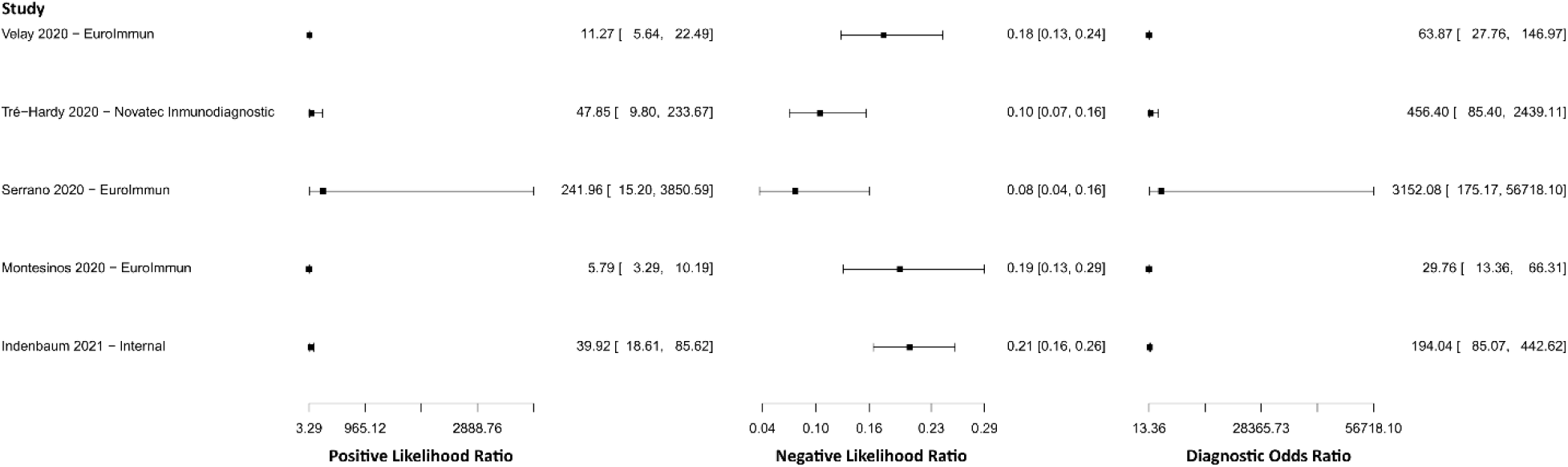
Study data and paired forest plot of the Positive Likelihood ratio, Negative likelihood ratio, and Diagnostic Odds ratio of enzyme-linked immunosorbent assay (ELISA) for IgA in the diagnosis of COVID-19. Positive Likelihood ratio, Negative likelihood ratio, and Diagnostic Odds ratio are reported with a mean (95% confidence limits). The Forest plot depicts the estimated sensitivity and specificity (black squares) and its 95% confidence limits (horizontal black line) [74,75,77,89,90].

**Supplementary 7.**
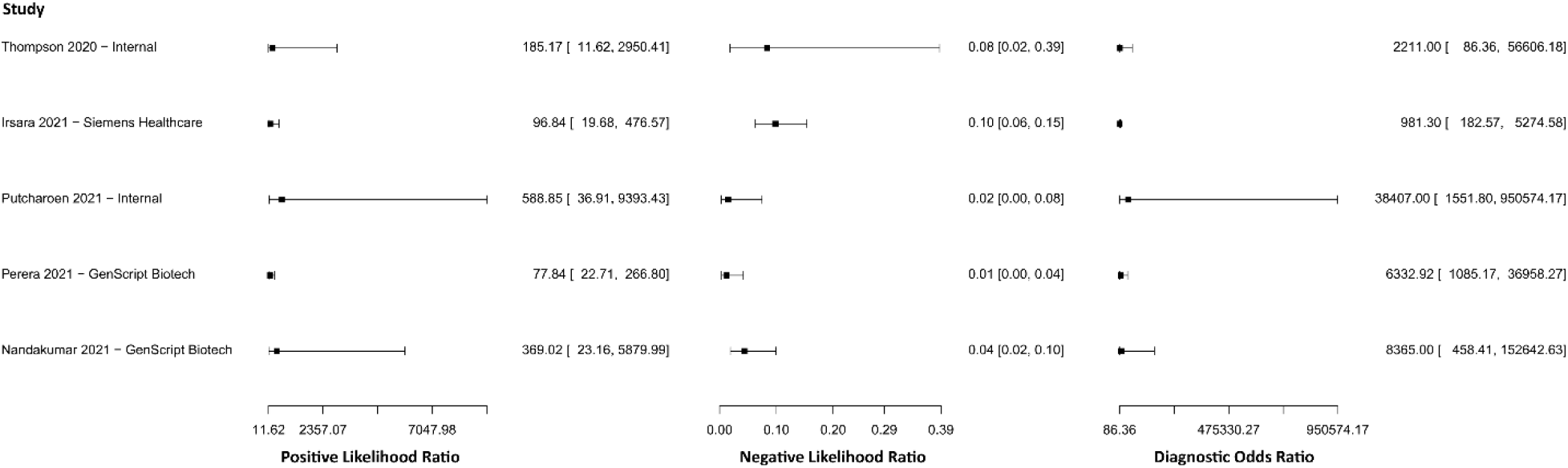
Study data and paired forest plot of the Positive Likelihood ratio, Negative likelihood ratio, and Diagnostic Odds ratio of antiviral neutralization bioassay (ANB) in the diagnosis of COVID-19. Positive Likelihood ratio, Negative likelihood ratio, and Diagnostic Odds ratio are reported with a mean (95% confidence limits). The Forest plot depicts the estimated sensitivity and specificity (black squares) and its 95% confidence limits (horizontal black line) [91–95].

**Supplementary 8.**
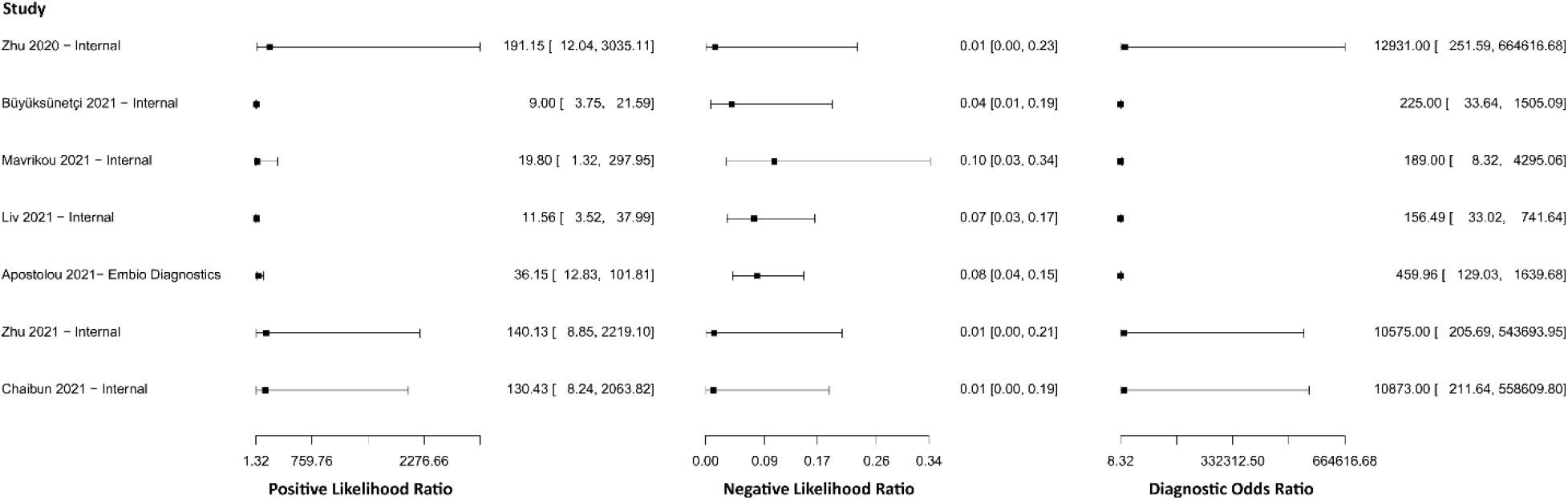
Study data and paired forest plot of the Positive Likelihood ratio, Negative likelihood ratio, and Diagnostic Odds ratio of biosensors (BS) in the diagnosis of COVID-19. Positive Likelihood ratio, Negative likelihood ratio, and Diagnostic Odds ratio are reported with a mean (95% confidence limits). The Forest plot depicts the estimated sensitivity and specificity (black squares) and its 95% confidence limits (horizontal black line) [96–102].

**Supplementary 9.**
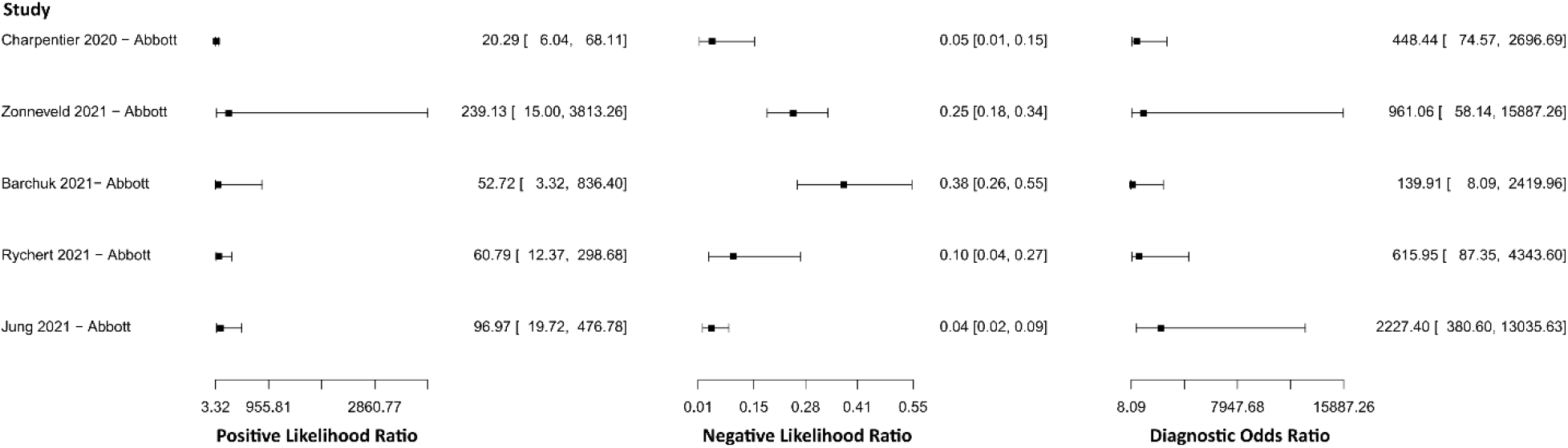
Study data and paired forest plot of the Positive Likelihood ratio, Negative likelihood ratio, and Diagnostic Odds ratio of chemiluminescence immunoassay (CLIA) for IgG in the diagnosis of COVID-19. Positive Likelihood ratio, Negative likelihood ratio, and Diagnostic Odds ratio are reported with a mean (95% confidence limits). The Forest plot depicts the estimated sensitivity and specificity (black squares) and its 95% confidence limits (horizontal black line) [76,77,79,88,103–107].

**Supplementary 10.**
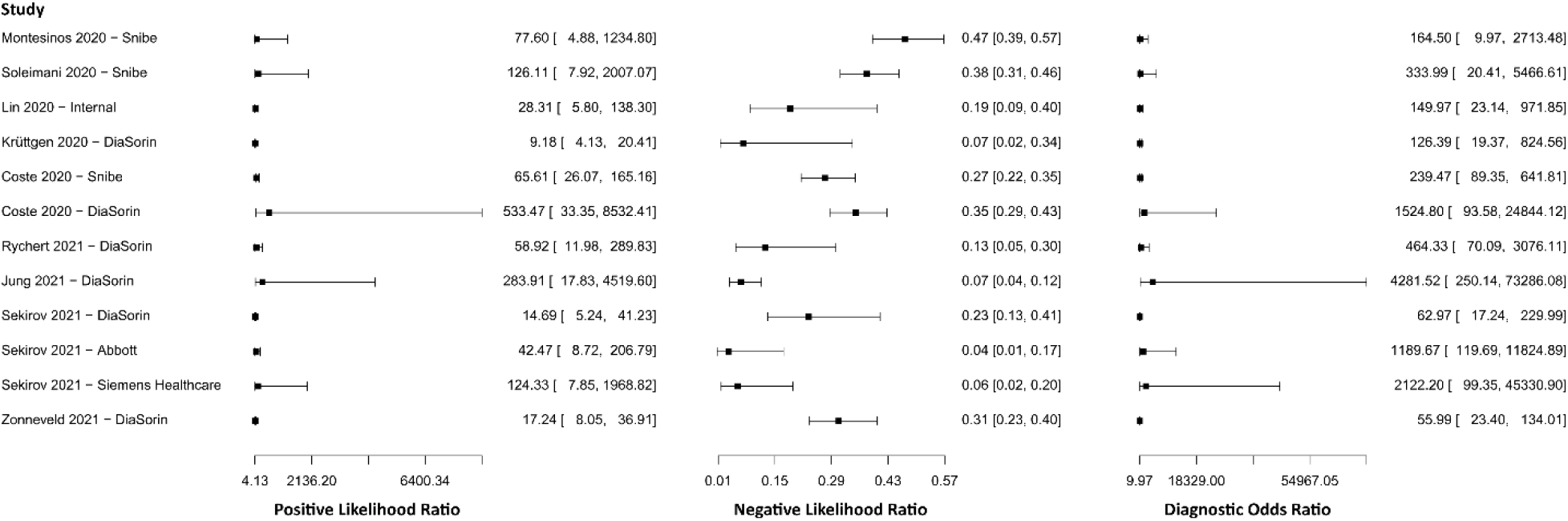
Study data and paired forest plot of the Positive Likelihood ratio, Negative likelihood ratio, and Diagnostic Odds ratio of chemiluminescence immunoassay (CLIA) for IgM in the diagnosis of COVID-19. Positive Likelihood ratio, Negative likelihood ratio, and Diagnostic Odds ratio are reported with a mean (95% confidence limits). The Forest plot depicts the estimated sensitivity and specificity (black squares) and its 95% confidence limits (horizontal black line) [77,79,103,104,106].

**Supplementary 11.**
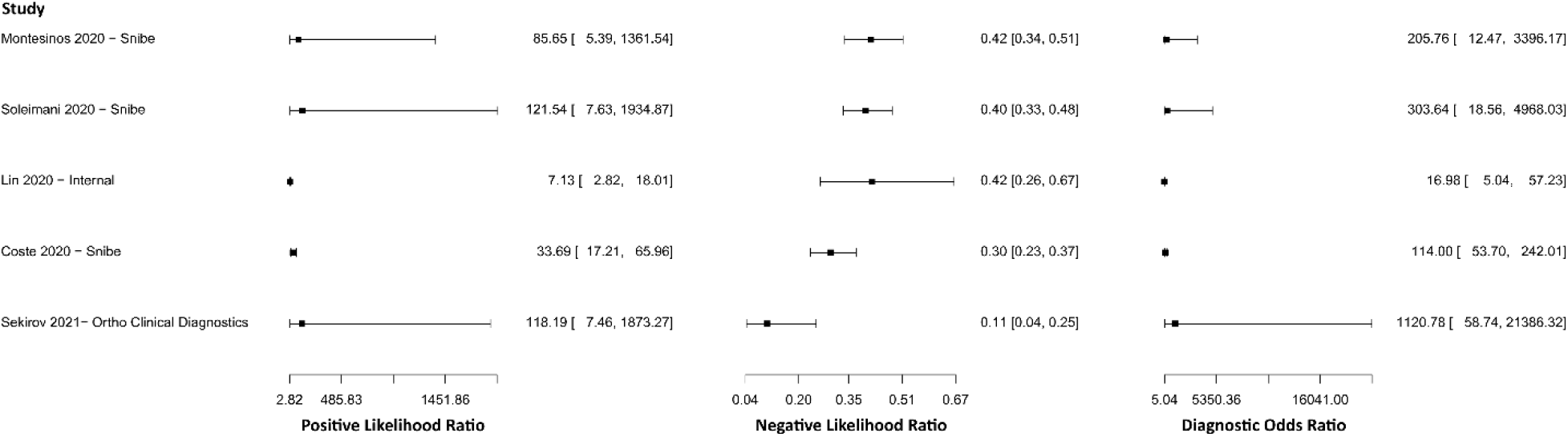
Study data and paired forest plot of the Positive Likelihood ratio, Negative likelihood ratio, and Diagnostic Odds ratio of chemiluminescence immunoassay (CLIA) for IgM-IgG in the diagnosis of COVID-19. Positive Likelihood ratio, Negative likelihood ratio, and Diagnostic Odds ratio are reported with a mean (95% confidence limits). The Forest plot depicts the estimated sensitivity and specificity (black squares) and its 95% confidence limits (horizontal black line) [77,85,103,106,108].

**Supplementary 12.**
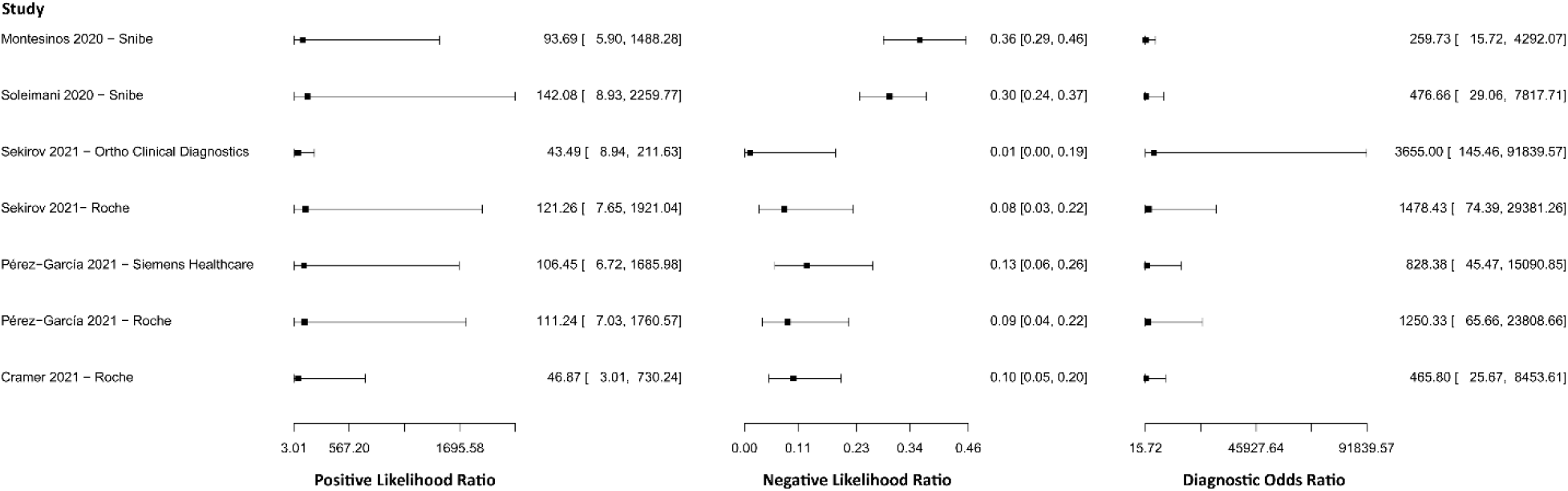
Study data and paired forest plot of the Positive Likelihood ratio, Negative likelihood ratio, and Diagnostic Odds ratio of lateral flow immunoassay (LFIA) for IgG in the diagnosis of COVID-19. Positive Likelihood ratio, Negative likelihood ratio, and Diagnostic Odds ratio are reported with a mean (95% confidence limits). The Forest plot depicts the estimated sensitivity and specificity (black squares) and its 95% confidence limits (horizontal black line) [75,77,79,109–116].

**Supplementary 13.**
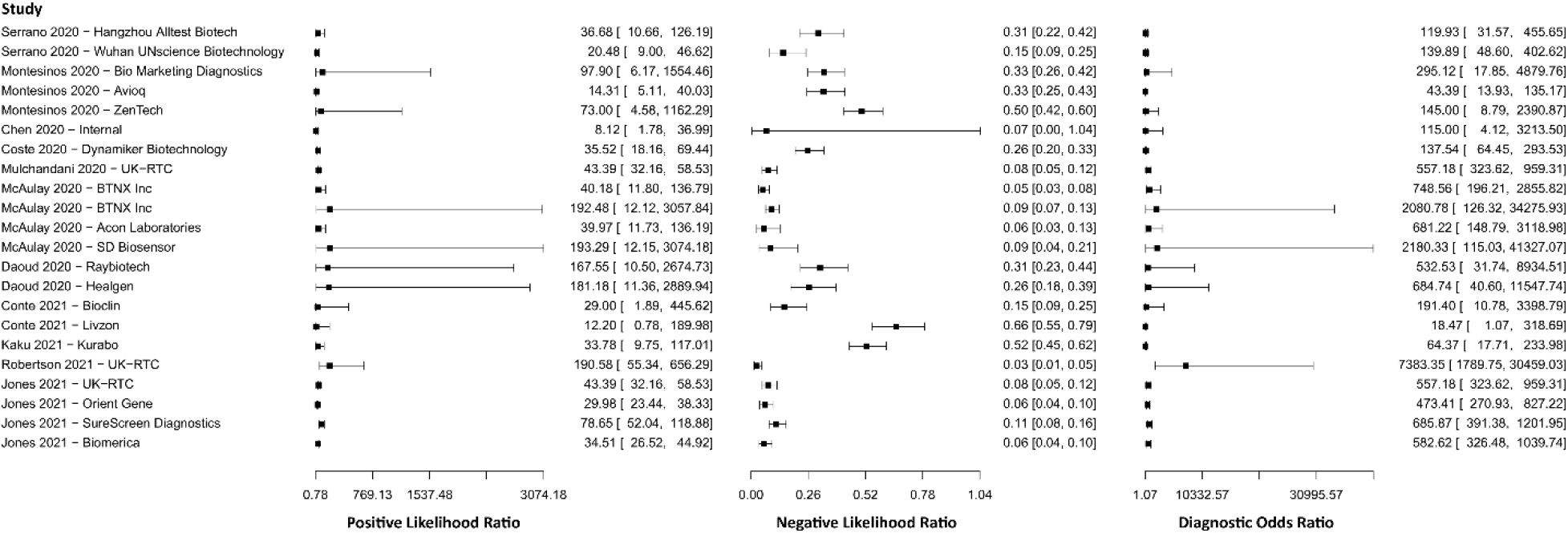
Study data and paired forest plot of the Positive Likelihood ratio, Negative likelihood ratio, and Diagnostic Odds ratio of lateral flow immunoassay (LFIA) for IgM in the diagnosis of COVID-19. Positive Likelihood ratio, Negative likelihood ratio, and Diagnostic Odds ratio are reported with a mean (95% confidence limits). The Forest plot depicts the estimated sensitivity and specificity (black squares) and its 95% confidence limits (horizontal black line) [75,77,79,111,112,115].

**Supplementary 14.**
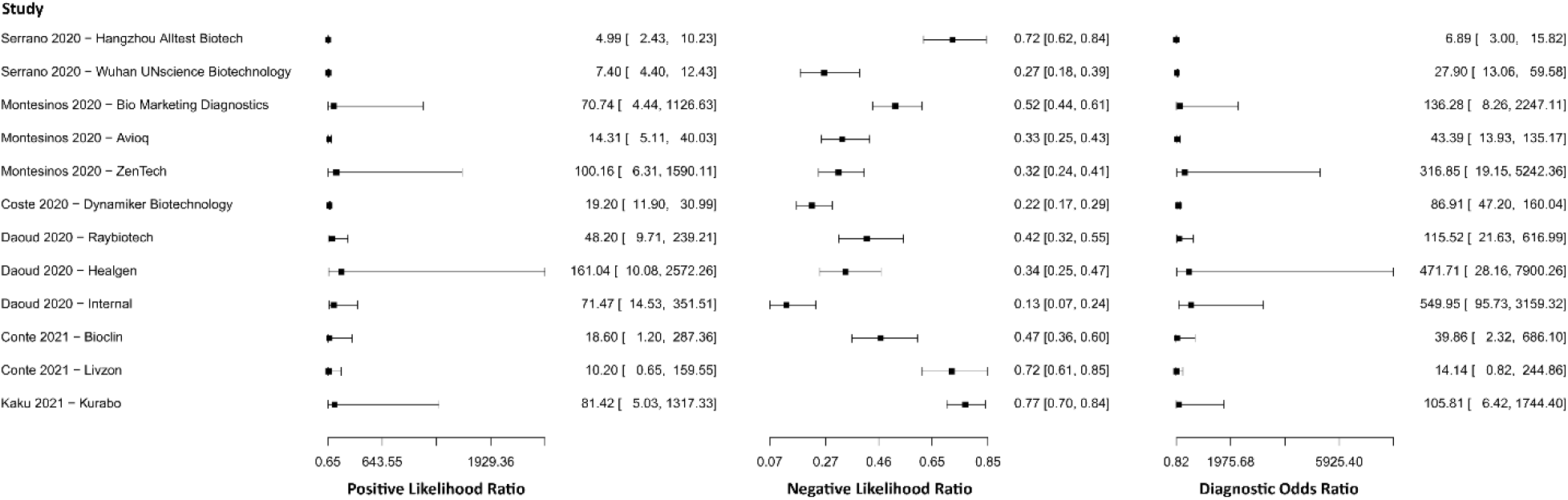
Study data and paired forest plot of the Positive Likelihood ratio, Negative likelihood ratio, and Diagnostic Odds ratio of lateral flow immunoassay (LFIA) for IgM-IgG in the diagnosis of COVID-19. Positive Likelihood ratio, Negative likelihood ratio, and Diagnostic Odds ratio are reported with a mean (95% confidence limits). The Forest plot depicts the estimated sensitivity and specificity (black squares) and its 95% confidence limits (horizontal black line) [75,77,85,90,108,109,115,117,118].

**Supplementary 15.**
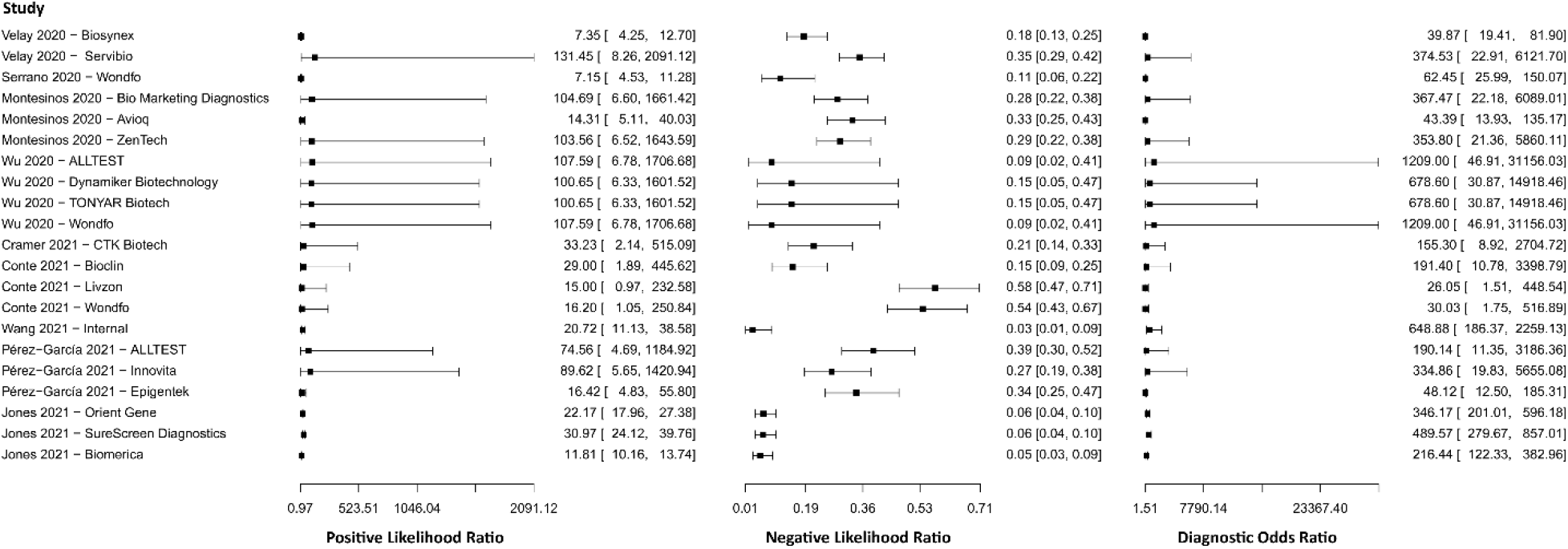
Study data and paired forest plot of the Positive Likelihood ratio, Negative likelihood ratio, and Diagnostic Odds ratio of lateral flow immunoassay (LFIA) for N-protein in the diagnosis of COVID-19. Positive Likelihood ratio, Negative likelihood ratio, and Diagnostic Odds ratio are reported with a mean (95% confidence limits). The Forest plot depicts the estimated sensitivity and specificity (black squares) and its 95% confidence limits (horizontal black line) [119–132].

**Supplementary 16.**
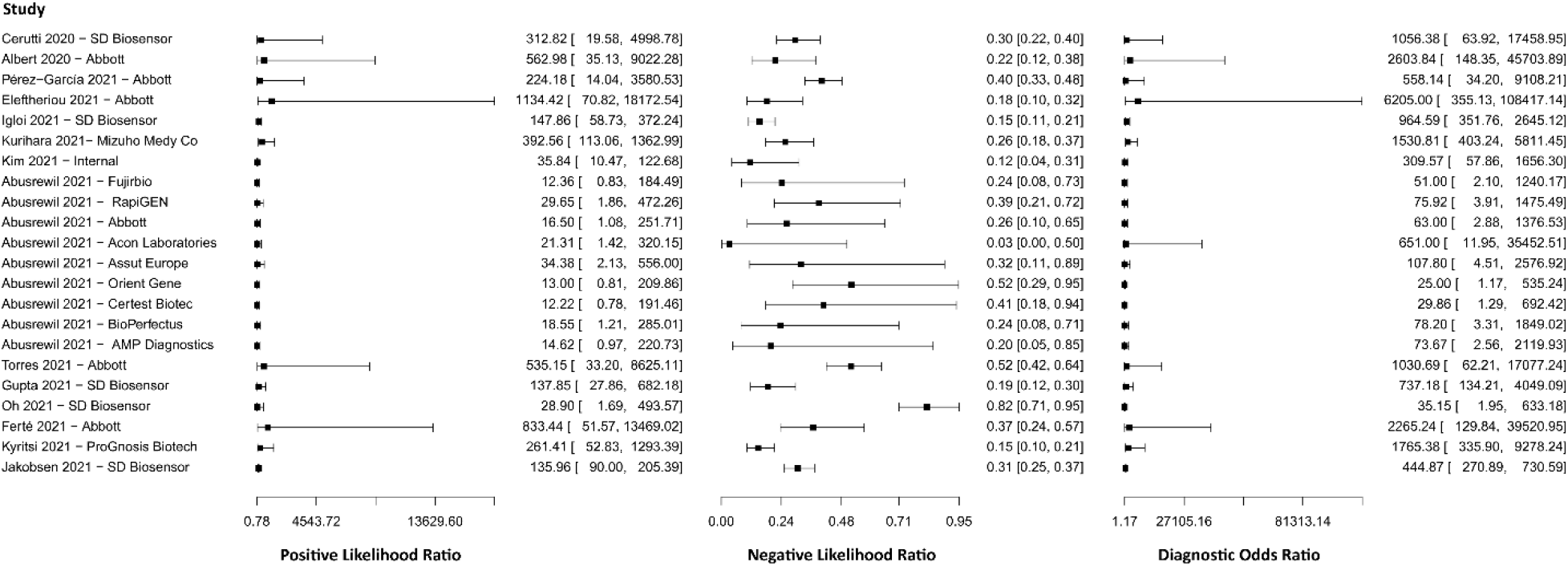
Study data and paired forest plot of the Positive Likelihood ratio, Negative likelihood ratio, and Diagnostic Odds ratio of chemiluminescent microparticle immunoassay (CMIA) in the diagnosis of COVID-19. Positive Likelihood ratio, Negative likelihood ratio, and Diagnostic Odds ratio are reported with a mean (95% confidence limits). The Forest plot depicts the estimated sensitivity and specificity (black squares) and its 95% confidence limits (horizontal black line) [81,88,105,107,133].

**Supplementary 17.**
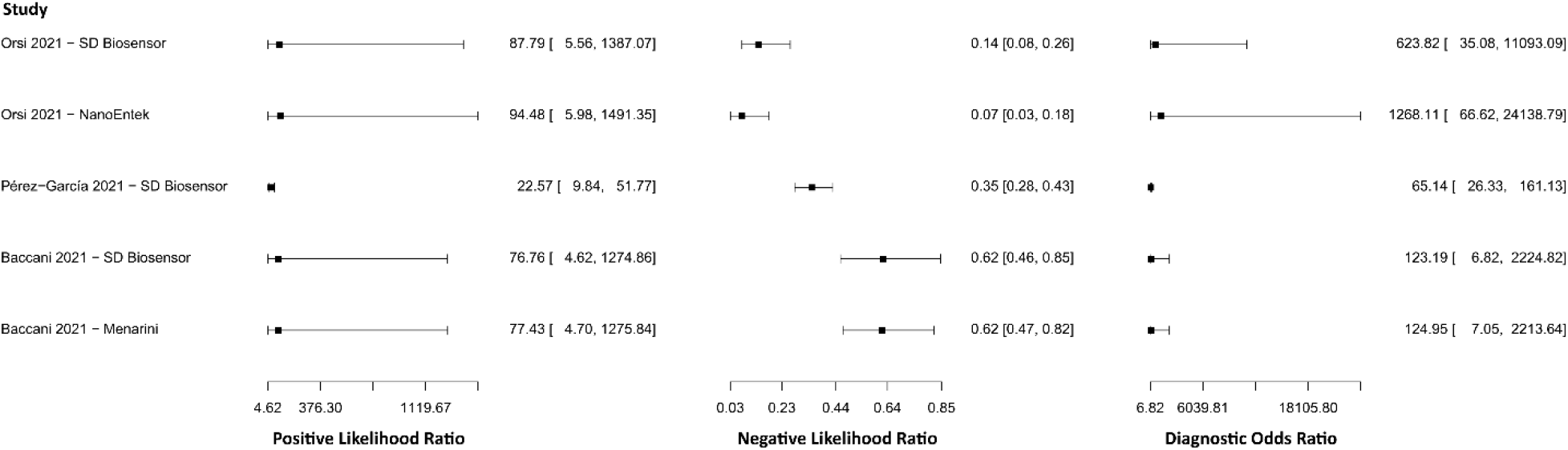
Study data and paired forest plot of the Positive Likelihood ratio, Negative likelihood ratio, and Diagnostic Odds ratio of fluorescence immunoassay (FIA) in the diagnosis of COVID-19. Positive Likelihood ratio, Negative likelihood ratio, and Diagnostic Odds ratio are reported with a mean (95% confidence limits). The Forest plot depicts the estimated sensitivity and specificity (black squares) and its 95% confidence limits (horizontal black line) [121,134,135].

**Table S1.**
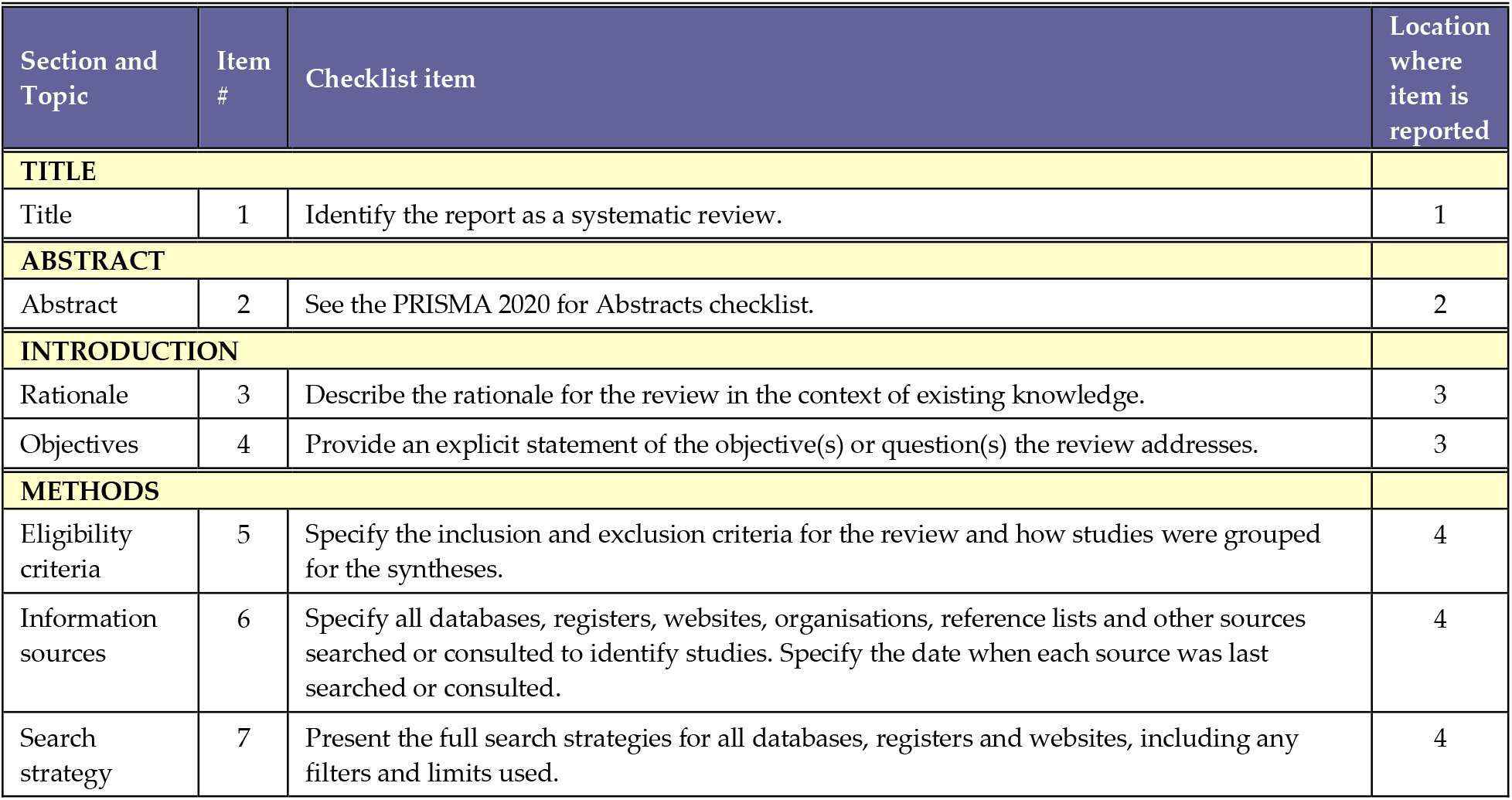
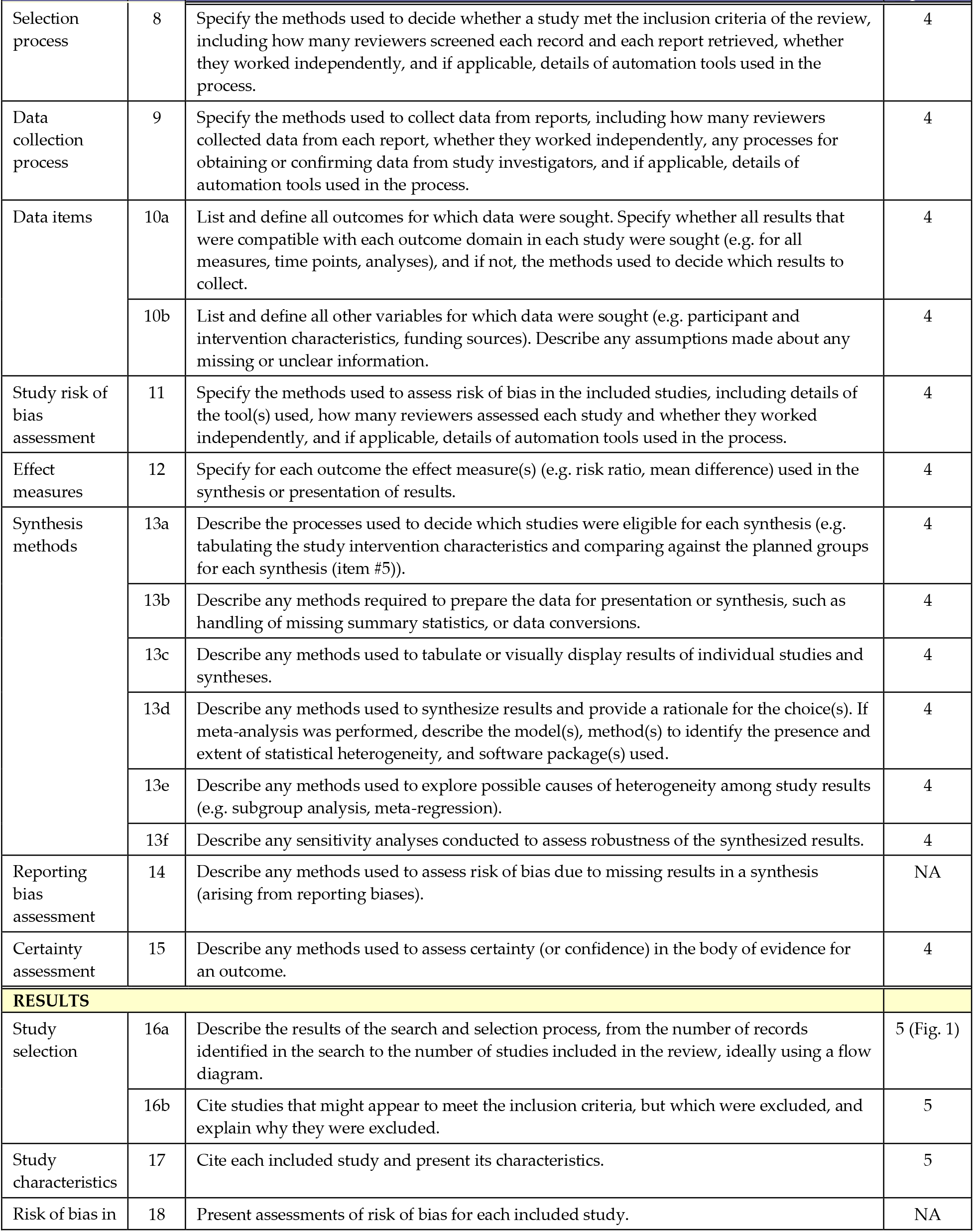
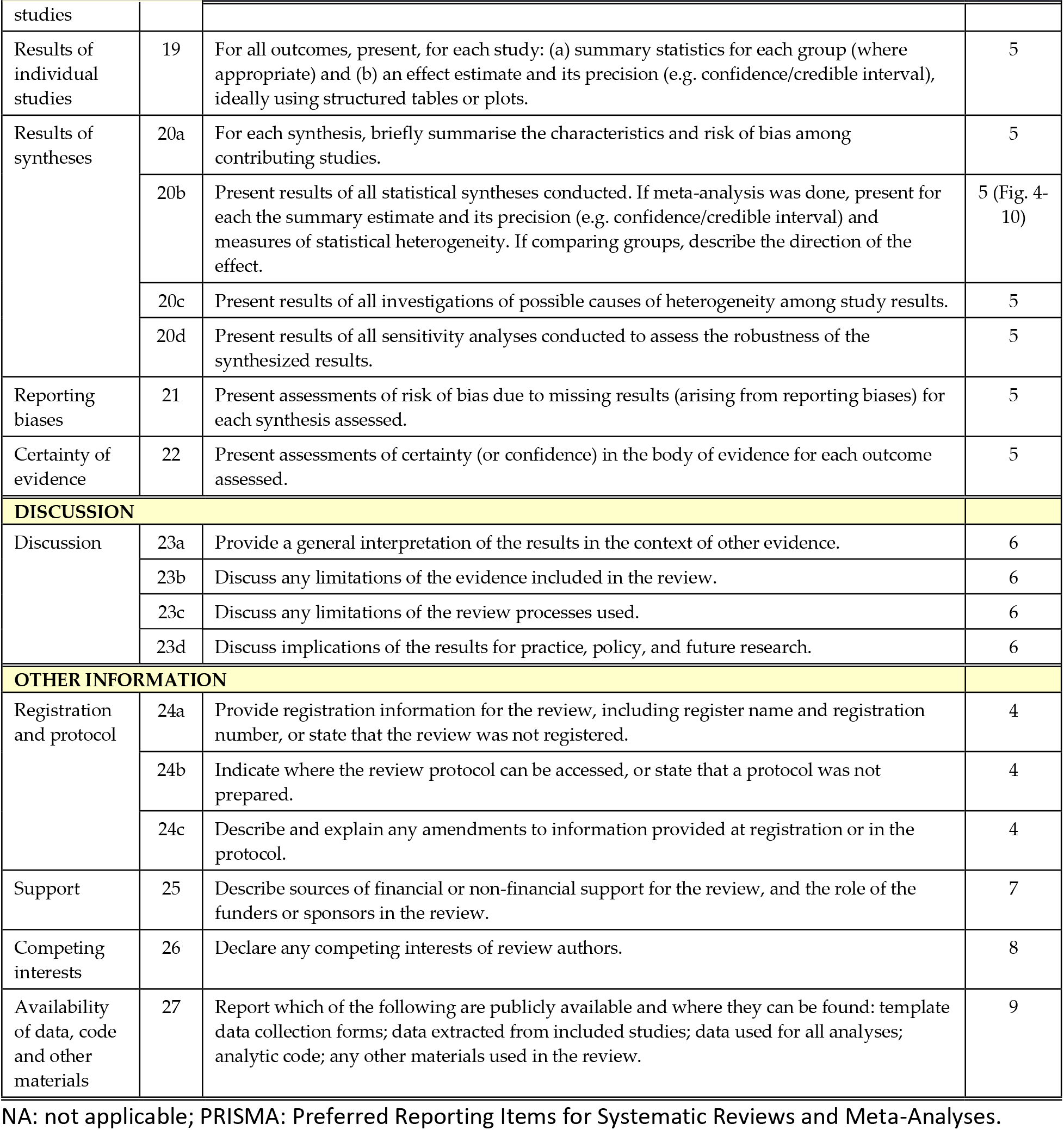
PRISMA 2020 Checklist.

